# Endemic-epidemic modelling of school closure to prevent spread of COVID-19 in Switzerland

**DOI:** 10.1101/2023.03.21.23287519

**Authors:** M. Bekker-Nielsen Dunbar, F. Hofmann, S. Meyer, L. Held

## Abstract

The coronavirus disease 2019 (COVID-19) pandemic disrupted daily life and changes to routines were made in accordance with public health regulations. In 2020, nonpharmaceutical interventions were put in place to reduce exposure to and spread of the disease. The goal of this work was to quantify the effect of school closure during the first year of COVID-19 pandemic in Switzerland. This allowed us to determine the usefulness of school closures as a pandemic countermeasure for emerging coronaviruses in the absence of pharmaceutical interventions. The use of multivariate endemic-epidemic modelling enabled us to analyse disease spread between age groups which we believe is a necessary inclusion in any model seeking to achieve our goal. Sophisticated time-varying contact matrices encapsulating four different contact settings were included in our complex statistical modelling approach to reflect the amount of school closure in place on a given day. Using the model, we projected case counts under various transmission scenarios (driven by implemented social distancing policies). We compared these counterfactual scenarios against the true levels of social distancing policies implemented, where schools closed in the spring and reopened in the autumn. We found that if schools had been kept open, the vast majority of additional cases would be expected among primary school-aged children with a small fraction of cases percolating into other age groups following the contact matrix structure. Under this scenario where schools were kept open, the cases were highly concentrated among the youngest age group. In the scenario where schools had remained closed, most reduction would also be expected in the lowest age group with less effects seen in other groups.

## 1 Introduction

It is known that school closures have an effect on social mixing and so school closures are considered useful for some infectious disease outbreaks but not necessarily all [Cowling et al., 2008]. The implications of school closures are manifold and are not restricted to changes in numbers of cases (knock-on effects include decreased socialisation skills among children and economic impacts through the reduced labour of guardians having to shift their focus to child rearing) meaning it is not a policy decision to be made lightly. As not everyone in a population attends school, we need age-stratified surveillance data to answer the question of what the impact of school closure is. In this work, we wish to determine the impact of school closures for COVID-19 control in Switzerland though the methods are applicable to other countries.

In earlier work [Bekker-Nielsen Dunbar et al., 2022] we considered evidence which suggested the effect of school closure in the canton of Zurich (in terms of reduction in disease transmission observed through a decrease in cases) seemed to not be large for the early coronavirus outbreak. The canton of Zurich is the most populous region of Switzerland. The analysis of data from the canton of Zurich suffered from low numbers of observed cases in the youngest age group. This proof-of-concept study provided a starting ground for further developing the methods used to examine these kinds of policy questions using endemic-epidemic models with time-varying weights. We now consider a longer time frame (until the end of 2020) and a greater population (the whole of Switzerland). This also allows us to evaluate the performance of the analysis at a greater resolution. Considering cases at national level rather than regional level induces additional challenges as social distancing policy varies across the country. As our study is not stratified by geographical region–our focus is age groups– these differences in policy need to be incorporated. Here we showcase how to incorpo-rate policy indicators which are more nuanced than those used in our previous work.

The endemic-epidemic framework for infectious disease modelling is a class of time-series based regression models used for the analysis of infectious disease case counts arising from routine surveillance systems. It is a versatile frameworkmodel which has been applied to the analysis of many disease outbreaks with varying characteristics. Endemic-epidemic modelling is considered a useful tool for emergency response related to infectious disease outbreaks as it fulfils many of the requirements for disaster response models raised by Brandeau et al. [2009]. In particular, endemic-epidemic modelling addresses real-world infectious disease problems such as detection of outbreaks and populations at increased risk and is designed for maximum usability by response planners by virtue of being released as open source publications and software which means we avoid issues with disease knowledge being pay-walled during ongoing outbreaks as seen in the 2014 Ebola virus disease outbreak [Dahn et al., 2015]. The framework also makes a good compromise between simplicity and complexity, and due to its statistical nature is designed in a manner which captures inherent uncertainties. Endemic-epidemic modelling facilitates knowing when disease is endemic (prevalence levels are in the range of expected values) and when disease is epidemic (incidence is higher than expected), at which point control measures may need introducing or intensifying. This work provides an insight into how control measures can be incorporated in endemic-epidemic models through the inclusion of time-varying contact matrices.

To accomplish our goal, we fit an endemic-epidemic model to a multivariate time series of age-stratified COVID-19 cases in Switzerland and then examine two counterfactual scenarios of the policy implemented; the true school closures consisted of schools being closed early in the year and reopening for the second half of the year (scenario A). We consider the counterfactual scenarios where schools did not close (al-ways open; scenario B and where the schools remain closed during the second half of the year (always closed; scenario C). The additional scenario is possible due to the longer time frame considered in this work. Scenario B is similar to the scenario considered in our earlier analysis.

## 2 Methods

This work is preregistered and has a study protocol [Bekker-Nielsen Dunbar et al., 2022] which is considered a useful manner of working but currently rare in epidemic modelling. The protocol outlines the modelling considerations we made before work began and may serve as a useful resource for the interested reader.

### 2.1 Data

In this work we consider daily data (*t* = 1, …, 312 = *T*) where the study period commences on 24^th^ February 2020 and the final observation at time *T* occurs on 31^st^ December 2020. COVID-19 case data is provided by the Swiss public health authority (Bundesamt für Gesundheit) and includes case counts by date reported stratified by age group. We asked for cases given by the same age groups we considered in our Zurich analysis as this roughly divides the population into those of compulsory education age (0–14 year olds are required to be in school when school is open), higher education and young workers (15–24), parents (25–44), middle-aged workers (45–65), retirees (66–79), and the elderly (80+). Our age group thresholds include the commonly used cut-off of 65 years of age considered in epidemiology, when health is expected to change.

Figure 1 shows the case data; the daily number of cases per 100, 000 age group population (upper panel), the distribution of cases per 100, 000 population over time (middle panel), and cases by weekday reported (lower panel). The upper panel shows the oscillatory behaviour known to epidemic curves as well as weekly systematic fluctuations in surveillance. The middle panel shows a shift in the age distribution of cases across the study period which further motivates the inclusion of age groups in our modelling approach. The lower panel shows the distribution of cases across the days of the week, where we see a systematic fluctuation in the reporting system. The legal workdays in Switzerland are Monday through Friday. Most cases are reported on Mondays which is the start of the week according to European norms and the number of reported cases drops across the week while fewer cases are reported on weekends (Saturday and Sunday).

**Figure 1:**
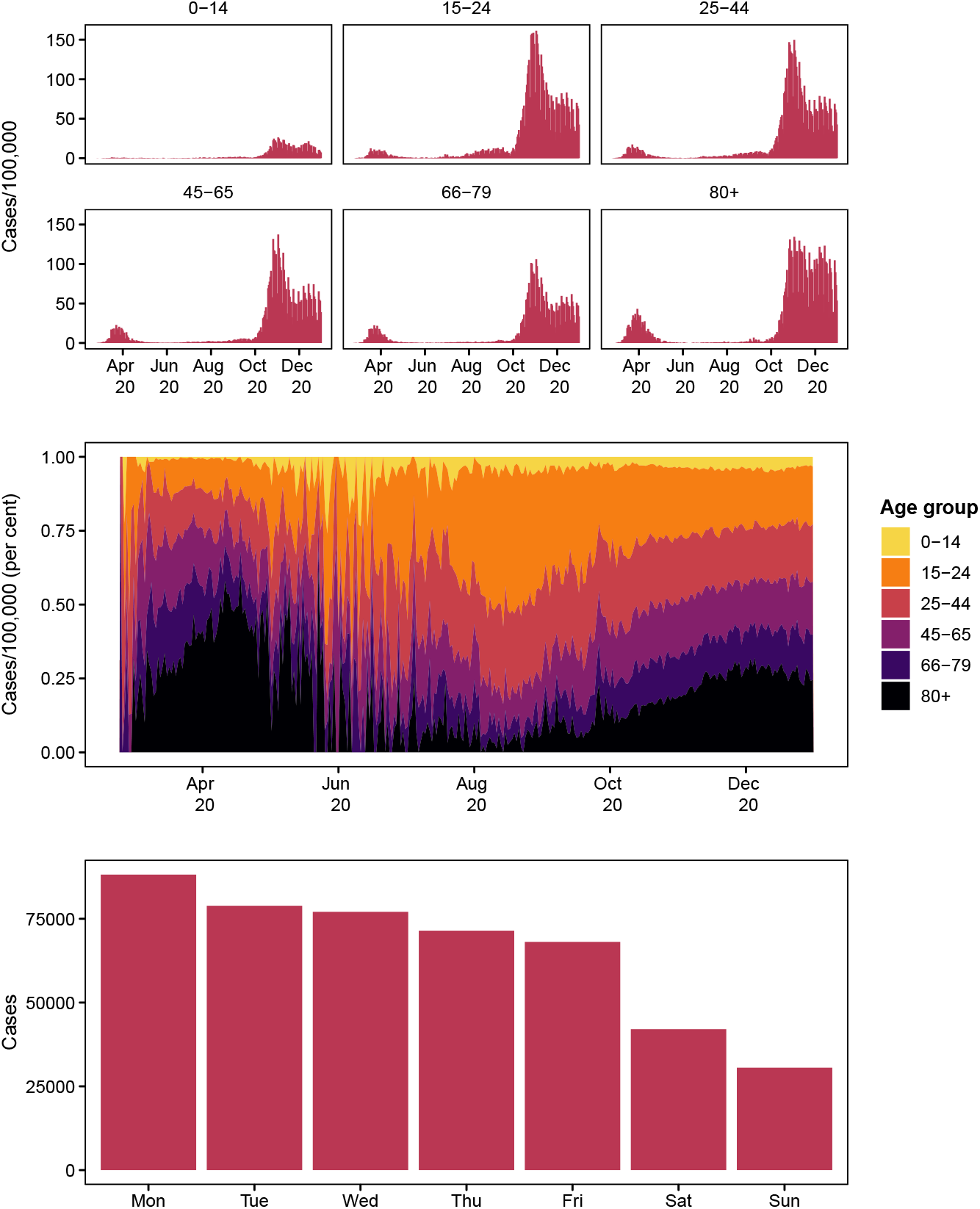
Daily COVID-19 cases per 100, 000 population (upper). Proportion of cases per 100, 000 population contributed by each age group (middle). Number of cases reported by weekday (lower)

To capture baseline transmission opportunities between age groups we include a contact matrix in our endemic-epidemic model. Contact matrices encapsulate the number of contacts an average person in the population has with other population members of the same and different ages in different settings such as the workplace or school. Their inclusion in the endemic-epidemic framework was an approach introduced by Meyer and Held [2017] which provided more realism than making an assumption of mixing patterns for the population. Existing contact diary-based contact matrices for Switzerland are only based on a small number of observations (54 observations) [Hoang et al., 2019], which led us to use a synthetic contact matrix in place of this empirical one. A synthetic contact matrix is constructed on the basis of demographic information. We chose to use the synthetic matrices by Mistry et al. [2021] as they were both 1) newer and 2) given with a certain level of uncertainty which we could incorporate into our modelling approach. The synthetic contact matrix is constructed on the basis of household size, school enrolment records, and employment data [see Mistry et al., 2021, for details].

From the synthetic contact matrix we obtain the per capita frequency of contact *c*_*a,a′,s*_ in setting *s* (shown in Figure 2 which describes the pattern of mixing in setting *s*) and the numbers of contacts *d*_*s*_ in setting *s* for constructing contact matrices for respiratory disease, which are 4.11 for household setting, 11.41 for school setting, 8.07 for work setting, and 2.79 for general community setting (shown in Figure 3). This means school has the largest weight and so changes to these contacts are expected to have the biggest impact. These construction weights *d*_*s*_ are provided with standard errors. When constructing *c*_*a,a′*_ we used the Swiss population (Table 1) rather than the Zurich population which we considered in earlier work [see Bekker-Nielsen Dunbar et al., 2022, for the analysis of Zurich] such that the population used to weight the synthetic contact matrix was the one being studied. This means the contact matrix used in this work is not exactly the same as the one considered previously. The Mistry et al. [2021] contact matrices are created with respiratory diseases in mind where school closure is a first line of defence against disease outbreaks. The synthetic contact matrix was used to inform the time-varying transmission weights *w*_*a,a′,t*_ which determine the amount of transmission between age group *a* and age group *a*^*′*^ at time *t*, which is explained in more detail below.

**Figure 2:**
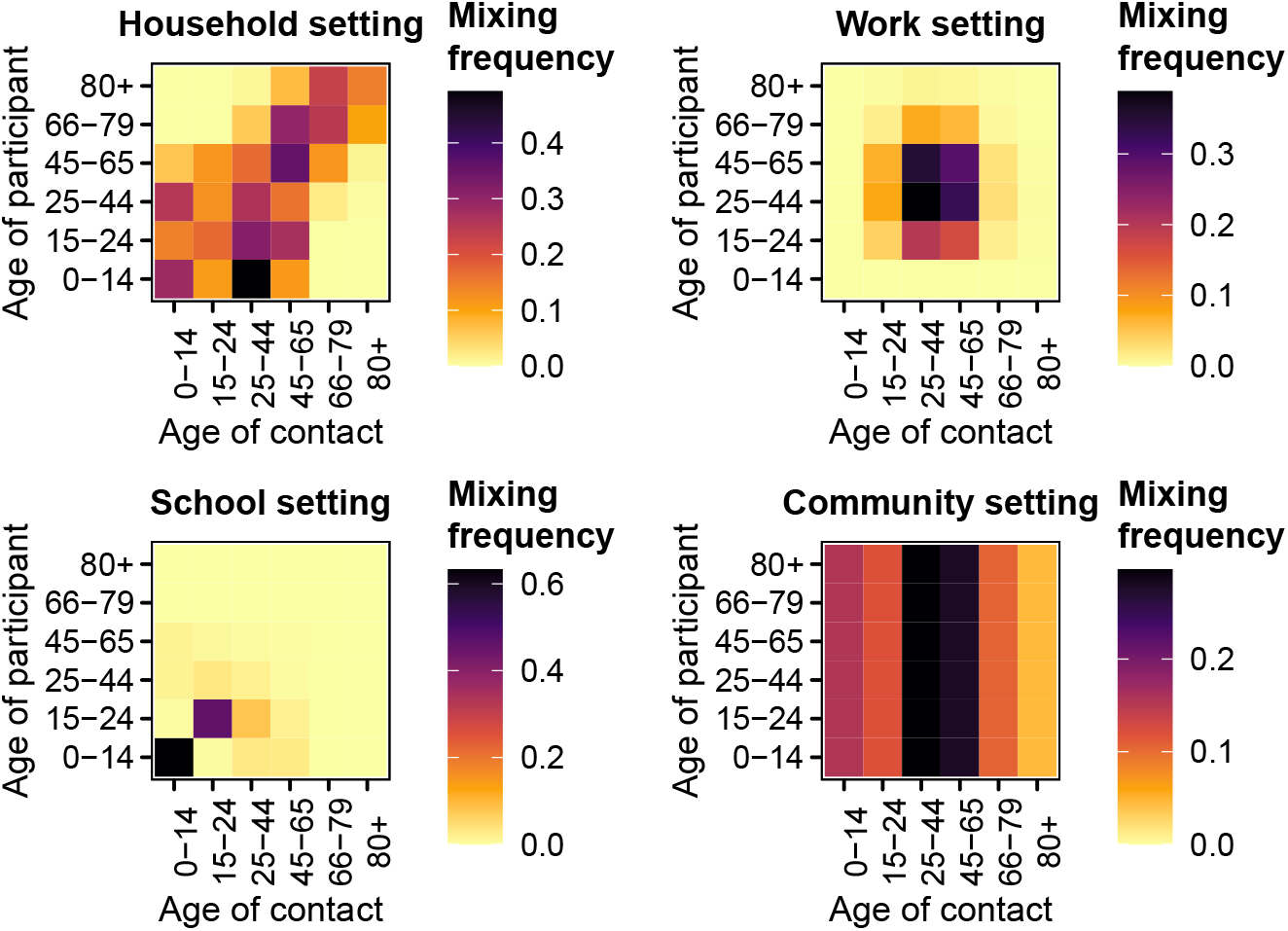
Unweighted contact matrices *c*_*a,a′,s*_ from Mistry et al. [2021] via Laboratory for the Modeling of Biological and Socio-technical Systems [2021]

**Figure 3:**
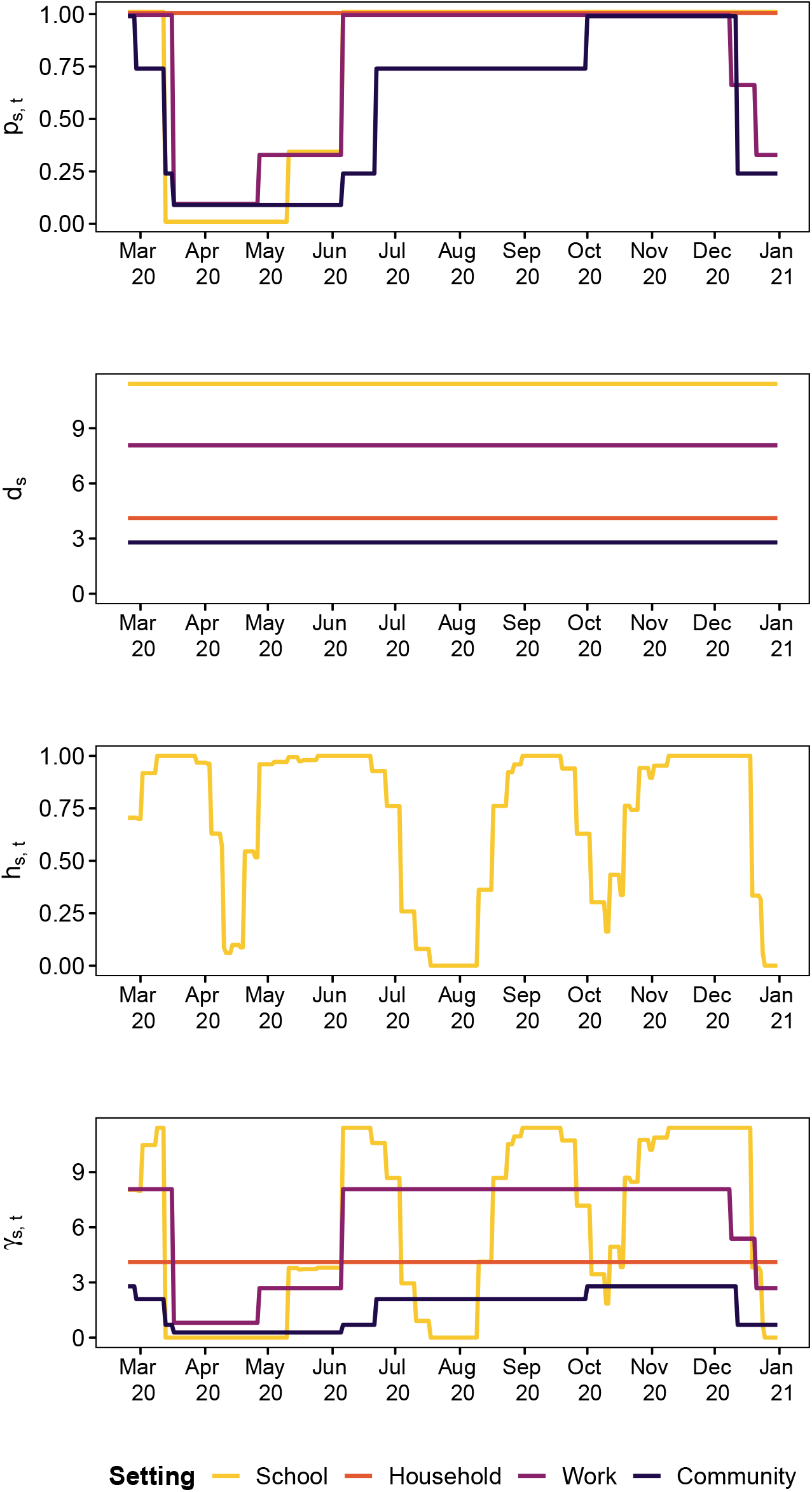
Time series of indicators (*p*_*s,t*_), disease setting-specific weights for contact matrices (*d*_*s*_), and holiday score (*h*_*s,t*_ for the school setting). The lowest panel shows the product, *γ*_*s,t*_

**Table 1:** Swiss population

| Age group | Population count | Proportion |
| --- | --- | --- |
| 0-14 | 1,294,918 | 0.150 |
| 15-24 | 901,783 | 0.105 |
| 25-44 | 2,383,179 | 0.277 |
| 45-65 | 2,511,163 | 0.292 |
| 66-79 | 1,061,320 | 0.123 |
| 80+ | 453,670 | 0.053 |

Contact setting-specific daily policy adjustments *p*_*s,t*_ are informed by information provided by the Swiss authorities. This information is used to quantify the amount of disease control measures enacted. We focus on those measures which have the aim to decrease contact. Following the Oxford University policy classifications we consider: school closure (“C1”), workplace closure (“C2”), and restrictions on gatherings (“C4”). We code our policy indicators to take the same levels as the Oxford scheme allowing researchers familiar with the controlled vocabulary established by that research group to comprehend our indicators. We reversed and rescaled the indicators such that *p*_*s,t*_ ∈ [0, 1] where 0 reflects a situation of maximal measures in place and 1 is full relaxation of measures (source information is in our study protocol https://osf.io/fgrdy).

The non-pandemic school closure adjustment *h*_*s,t*_ is created based on information from Schweizerische Konferenz der kantonalen Erziehungsdirektoren [2018] and reflects closures of school during the academic year due to half term and other school holidays. These closures reduce contact independently of disease control measures; notably Easter is a period where less contacts in school settings would be expected as Switzerland is a predominantly Christian country. The adjustment takes values *h*_*s,t*_ ∈ [0, 1] where 0 means that all schools in Switzerland are closed on that day and

1 means all schools are open. The values *h*_*s,t*_ takes for the school setting are shown in Figure 3 while *h*_*s,t*_ ≡ 1 for all other contact settings (meaning no adjustment). The calculation of *h*_*s,t*_ is informed by population data from Eurostat [Eurostat, 2021] (population by region). The construction of *p*_*s,t*_, *h*_*s,t*_, and *w*_*a,a′,t*_ is explained in more detail in the following section.

### 2.2 Model

In an endemic-epidemic model, case counts *Y*_*at*_ are indexed by time *t* and age group *a*. The age groups considered are 0–14, 15–24, 25–44, 45–65, 66–79, and 80+ years; the same used in Bekker-Nielsen Dunbar et al. [2022]. Case counts given past cases assumed to follow an overdispersed negative binomial distribution with age-dependent overdispersion parameters *ψ*_*a*_. The mean *λ*_*at*_ is additively decomposed into endemic and epidemic components. Log-linear predictors for the endemic and epidemic components are given by *ν*_*at*_ and *ϕ*_*at*_ respectively. The endemic component is additionally weighted by population fractions *e*_*a*_, where we used population data from Eurostat [2021] (population by age group), given in Table 1 to inform this part of the model.

The epidemic component is an autoregressive process driven by cases in other age groups *a*^*′*^ in previous time periods *t* − *d* up to a maximum lag of *d*_max_ where *u*_*d*_ determines by how much previous cases are weighted. We chose to use a Poisson-distributed lag distribution *u*_*d*_ such that the majority of the weight need not be given to the immediately preceeding cases allowing for a serial interval of more than a single day. The maximum lag represents the maximum length of the serial interval we might conceive in our modelling efforts; we chose *d*_max_ = 7 as the literature suggests early types of COVID-19 have a serial interval within a week.

Our endemic-epidemic model with time dependent [Bekker-Nielsen Dunbar et al., 2022, Grimée et al., 2021] contact matrix weights [Meyer and Held, 2017] and higher order lag [Bracher and Held, 2022] is given by

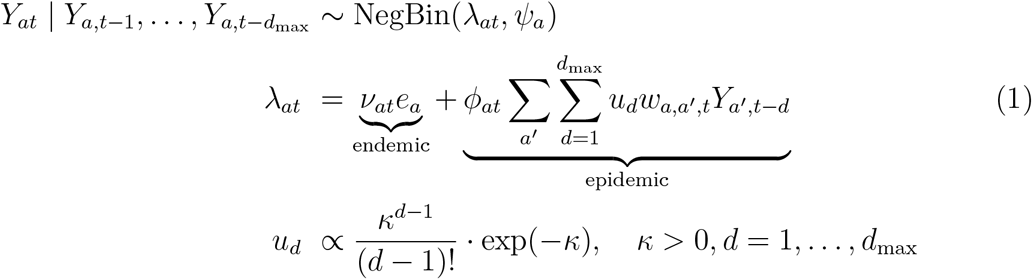

Transmission between age groups is determined by a time-dependent contact matrix *w*_*a,a′,t*_. The time-varying contact matrix *w*_*a,a′,t*_ is the total average contacts at time *t* constructed by a weighted sum

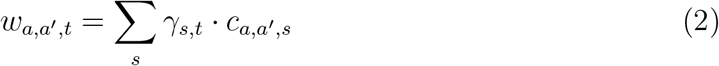

where *γ*_*s,t*_ is a weight that depends on the setting *s* the contact occurred in and changes occuring at time *t*. It is created from the combination of the weights used to construct contact matrices (*d*_*s*_ given by Mistry et al. [2021] which depend on setting *s*), the time-dependent setting-specific policy adjustments (*p*_*s,t*_ which depend on time *t* and setting *s*) and whether an adjustment needs to be made to incorporate nonpandemic school closure due to school holidays (*h*_*s,t*_ which depends on setting *s* as it only affects schools and time *t*):

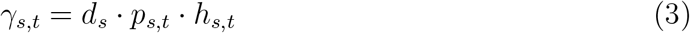

Since school holidays in Switzerland vary not only between regions, but also within them, we construct binary indicators for all of the sub-regions *r* within a region *R* where we assign 1 to a specific day *t* if *t* is not a school holiday and 0 otherwise. In a second step, we average the binary indicators of all sub-regions *r* within a region *R* in order to obtain a regional average indicator for that day. Subsequently, we use population weights to calculate the national indicator *h*_*s,t*_. The sub-regions are unweighted in our averaging as we were not able to determine population sizes at school district level. We calculate

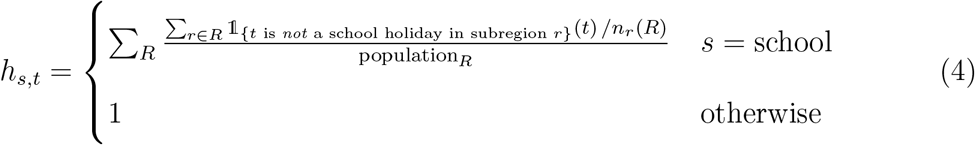

where 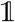 is an indicator function and *n*_*r*_(*R*) denotes the number of sub-regions within region *R*. This gives us a population-weighted indicator with values *h*_*s,t*_ ∈ [0, 1] which incorporates the variation of number of school children in regions.

We fit the model (1) with predictors

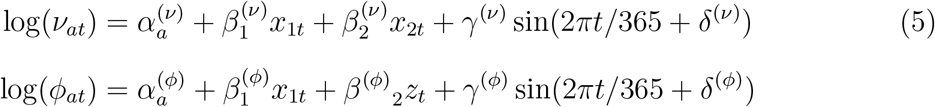

where *α*_*a*_ denotes a fixed effect of age group *a, x*_1*t*_ is an indicator for public holidays, *x*_2*t*_ is an indicator for weekends, and *z*_*t*_ are effect-coded weekday effects with Monday as the reference value (six in total). Effect-coded variables are also known as sum-to-zero contrasts. This means Monday always takes the value −1 and the weekday of interest takes the value 1 while all other weekdays are 0. We include a nonlinear time trend in the form of a sine-cosine wave expressed by its amplitude *γ* and phase *δ* [Paul et al., 2008]. Our model has 31 parameters (estimates are given in Table 2) which are estimated using a maximum likelihood approach computed with stan-dard errors. Information on the full model selection procedure (where we also considered effects of temperature, testing rate, and a linear time trend) can be found in the supporting information.

**Table 2:** Model parameter estimates

| Endemic |  |  | Epidemic |  |  | Other parameters |  |  |
| --- | --- | --- | --- | --- | --- | --- | --- | --- |
| Coefficient | Estimate | Std. Error | Coefficient | Estimate | Std. Error | Coefficient | Estimate | Std. Error |
| $\alpha_{0-14}^{(\nu)}$ | 2.806 | 0.218 | $\alpha_{0-14}^{(\phi)}$ | -3.776 | 0.059 | $\psi_{0-14}$ | 0.229 | 0.030 |
| $\alpha_{15-24}^{(\nu)}$ | 4.868 | 0.185 | $\alpha_{15-24}^{(\phi)}$ | -2.482 | 0.043 | $\psi_{15-24}$ | 0.125 | 0.013 |
| $\alpha_{25-44}^{(\nu)}$ | 4.036 | 0.185 | $\alpha_{25-44}^{(\phi)}$ | -2.172 | 0.027 | $\psi_{25-44}$ | 0.067 | 0.007 |
| $\alpha_{45-65}^{(\nu)}$ | 2.673 | 0.288 | $\alpha_{45-65}^{(\phi)}$ | -2.170 | 0.023 | $\psi_{45-65}$ | 0.059 | 0.006 |
| $\alpha_{66-79}^{(\nu)}$ | 2.246 | 0.321 | $\alpha_{66-79}^{(\phi)}$ | -1.883 | 0.027 | $\psi_{66-79}$ | 0.074 | 0.009 |
| | | | $\alpha_{80+}^{(\phi)}$ | -0.945 | 0.018 | $\psi_{80+}$ | 0.043 | 0.009 |
| | | | $\beta_{\text{day of the week Tuesday}}^{(\phi)}$ | 0.378 | 0.021 | | | |
| | | | $\beta_{\text{day of the week Wednesday}}^{(\phi)}$ | 0.119 | 0.022 | | | |
| | | | $\beta_{\text{day of the week Thursday}}^{(\phi)}$ | -0.032 | 0.022 | | | |
| | | | $\beta_{\text{day of the week Friday}}^{(\phi)}$ | 0.001 | 0.022 | | | |
| | | | $\beta_{\text{day of the week Saturday}}^{(\phi)}$ | -0.404 | 0.023 | | | |
| | | | $\beta_{\text{day of the week Sunday}}^{(\phi)}$ | -0.684 | 0.024 | | | |
| $\beta_{\text{weekend}}^{(\nu)}$ | -0.850 | 0.100 | | | | | | |
| $\beta_{\text{public holiday}}^{(\nu)}$ | -0.582 | 0.462 | $\beta_{\text{public holiday}}^{(\phi)}$ | -0.327 | 0.063 | | | |
| $\beta_{\text{amplitude}}^{(\nu)}$ | 2.070 | 0.191 | $\beta_{\text{amplitude}}^{(\phi)}$ | 0.711 | 0.018 | | | |
| $\beta_{\text{phase}}^{(\nu)}$ | -2.487 | 0.030 | $\beta_{\text{phase}}^{(\phi)}$ | 1.447 | 0.012 | | | |
| | | | | | | $\log \kappa$ | 0.082 | |

### 2.3 Counterfactual scenario prediction

Determining the expected size of the outbreak is crucial to policy makers who need to determine how resources are to be allocated. As the outbreak is ongoing, the predicted final size considered here is the predicted number of infections over the time window considered rather than the traditional metric considered by users of compartmental models: the total number of infections over the entire outbreak period. Predicted cases are based on a path trajectory (a long-term expected prediction calculated recursively on the basis of one-step predictions) following Held et al. [2017] assuming no changes to the model parameters across the scenarios considered. This means we predict the model (1) with the given *e*_*a*_ and fitted 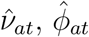, and *û*_*d*_ effects for three different versions of *w*_*a,a′,t*_ (for scenarios A, B, and C). The two counterfactual scenarios are implemented by including transmission weights informed by *γ*_*s,t,a,a′*_ = *d*_*s*_ · *q*_*s,t,a,a′*_ · *h*_*s,t*_ where *γ* now depends on age group. In particular we consider three scenarios (provided with the shorthand names we use based on their effect on the youngest age group):

Scenario A (“true measures”) This is the true measures scenario where schools closed in the spring and reopened in the summer where *w*_*a,a′,t*_ is populated by the relevant policy information without adjustment as in (2). This is the same scenario considered in model fitting to obtain the model coefficients used in prediction of final size and simulation of uncertainty for the prediction.

Scenario B (“schools open”) This is a scenario where schools are never closed for the youngest age group (0–14), i.e. remain open across the entire study period. All other measures are as in Scenario A.

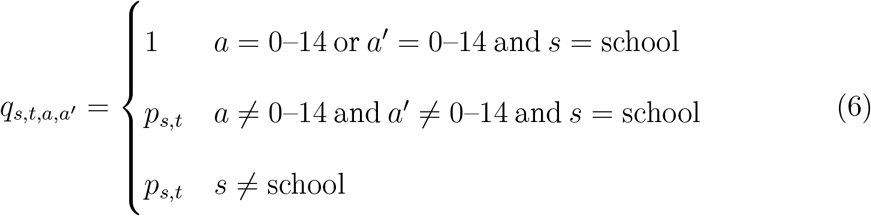

Scenario C (“schools closed”) This is a scenario where schools close and remain closed. School closure once again affects age group 0–14 and their contacts. All other measures are as in Scenario A. We implement this by setting

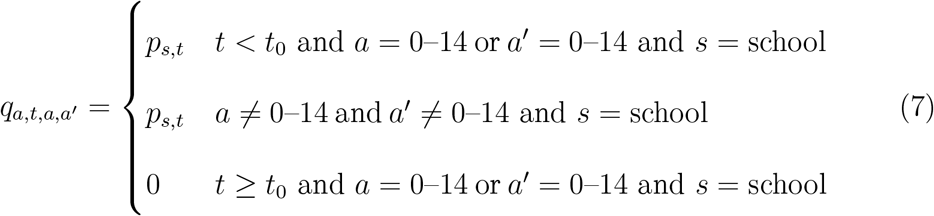

where *t*_0_ denotes the date schools are first closed (16^th^ March 2020).

The changes only affect age group 0–14 when the contact matrix is multiplied by *γ*_*s,t,a,a′*_ so the 0–14 row and column in Figure 2 are changed in the school setting (school aged children and their contacts). The time series of all four setting specific policy indicators *p*_*s,t*_ for the different scenarios can be seen in Figure **??** (the building blocks of (2), (6), (7)). The truth (scenario A) is expected to lie somewhere between the two counterfactual scenarios (scenarios B and C). Examining the deviation these scenarios have allows use to evaluate the effect of disease control measures used. It is implicitly assumed that the fitted effects 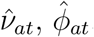, *û*_*d*_ do not vary across scenarios.

To incorporate parameter uncertainty in our projections, we utilise Monte Carlo simulation. We sample the weights *d*_*s*_ with uncertainty estimates given in Mistry et al. [2021] assuming they are independently normally distributed. To incorporate model uncertainty we sample the coefficients 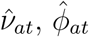 of our fitted endemic-epidemic model assuming a multivariate normal distribution; the asymptomatic normal distribution of the maximum-likelihood estimates. Using these *n* = 1000 samples we then use the path trajectory prediction approach to obtain *n* simulated expected case counts under the scenarios considered. This enables us to incorporate uncertainty in our projections. We examine the expected increase in cases when schools are always open (scenario B) and the expected decrease in cases when schools are always closed (scenario C) and compare this with the expected number of cases under the policy used (scenario A).

We also conducted sensitivity analyses of the assumptions made in constructing the transmission weights *w*_*a,a′,t*_. The sensitivity analyses attempt to provide further realism with respect to how household contacts may be affected by school closure. This provides additional extensions to the analysis of Zurich data as here we only considered household contacts to not be affected by policy *p*_*s,t*_|_household_ ≡ 1. The sensitivity analyses can be found in the supporting information.

## 3 Results

In total 256 models were fit to the outbreak data and Bayesian information criterion was used as a goodness-of-fit measure to determine the best fitting model (see the supporting information for details). We chose this as Bayesian information criterion should fit the correct model in theory while Akaike information criterion would be expected to overfit. Due to diverging estimates in the model–likely due to low values in the transmission weights matrix *w*_*a,a′,t*_ or low case counts *Y*_*at*_ observed in certain age groups–models which did not have converging effects were excluded from the selection process. Divergence was determined on the basis of the size of the standard deviation of the estimated model coefficients. It may happen that the additive decomposition into endemic and epidemic components is not identifiable.

In particular, 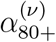 (the fixed effect of the oldest age group in the endemic component) was excluded due to having a very small estimate with a huge standard error. This means the coefficient 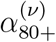 was restricted to be zero on the log-scale while the corresponding epidemic effect 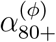 was estimated from the data. The best fitting model has 31 parameters including the lag parameter *κ*. The model contains systematic fluctuations in the form of weekly effects, as we expected based on the exploration of the case data, and additional fluctuations in the form of the sine-cosine waves. As we only use one year’s worth of data in this work, we cannot denote this fluctuating trend “seasonality” but with a longer time frame it would be expected to capture such effects. There is not much knowledge about seasonal variation of COVID-19 at the time of analysis so we note that with only one harmonic in the epidemic component and not even a whole year of data, this may induce additional uncertainty in our simulations and predictions.

### 3.1 Model fit

The model has a good fit to the case data based on visual inspection (Figure 4 upper panel). The serial interval peaks somewhat early (Figure 4 lower panel) compared with what is expected from the literature. This has been observed in other endemic-epidemic models for COVID-19 and is thus likely an artefact of the model. The model estimates fewer cases on public holidays and weekends, which aligns with our intuition based on the exploratory data analysis of the case counts.

**Figure 4:**
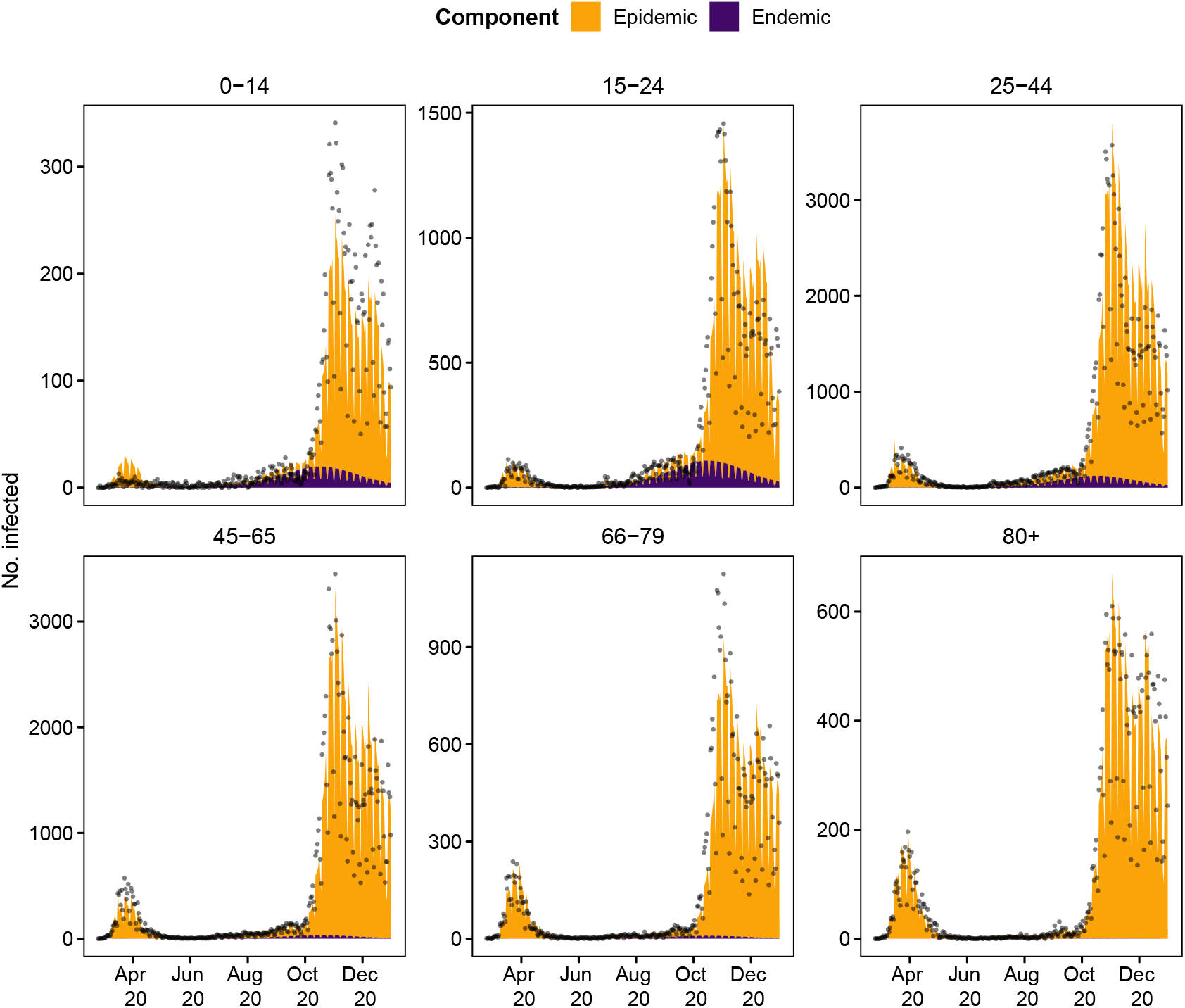
Model fit and cases

There are much greater effects of age in the endemic component, exp(*α*^(*ν*)^) ranges from 9.450 to 130.061 (and *α*^(*ν*)^ ≡ 1) compared with the epidemic component where exp(*α*^(*ϕ*)^) are all between 0.023 and 0.389. The fixed effect of age in the epidemic component which smaller in size shows a more obvious pattern of age dependency; being older makes a case more likely. The effect of day of the week is greater for all exp(*β*^(*ϕ*)^) compared with the weekend effect exp(*β*^(*ν*)^). However Friday seems to have less average autocorrelation. Public holidays have a greater effect on cases in the epidemic component based on the estimated effect size; this could reflect changes in contact patterns hence transmission opportunities on those days. The sine-cosine wave takes greater amplitude values in the endemic component, meaning the wave has stronger variation (relative to the baseline). The greatest value of the overdispersion (excess variance) parameter *ψ*_*a*_ is found the for the youngest age group 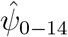 while the small-est value is found for the oldest age group 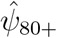.

### 3.2 Disease control scenarios

The path trajectories allow us to examine temporal changes that are not evident when projections are summarised as a final size estimate. The counterfactual scenarios’ path trajectories are compared in Figure 5 which shows the ratio of predicted cases under a counterfactual scenario and predicted cases under the original scenario A for the 90 days after measures were introduced (scenario B) or lifted (scenario C). This means we conditioned on fewer days when predicting for scenario B and had higher case counts included in our prediction of scenario C (as April was included). Large temporal differences are not found in the final epidemic size estimates (Figure 6). Scenarios A and C differ with regards to when schools are closed. They show much more distinct behaviour for school-aged children. We see the difference in patterns for age group 0–14 seems to be correlated with school holidays *h*_*s,t*_ (Figure 3).

**Figure 5:**
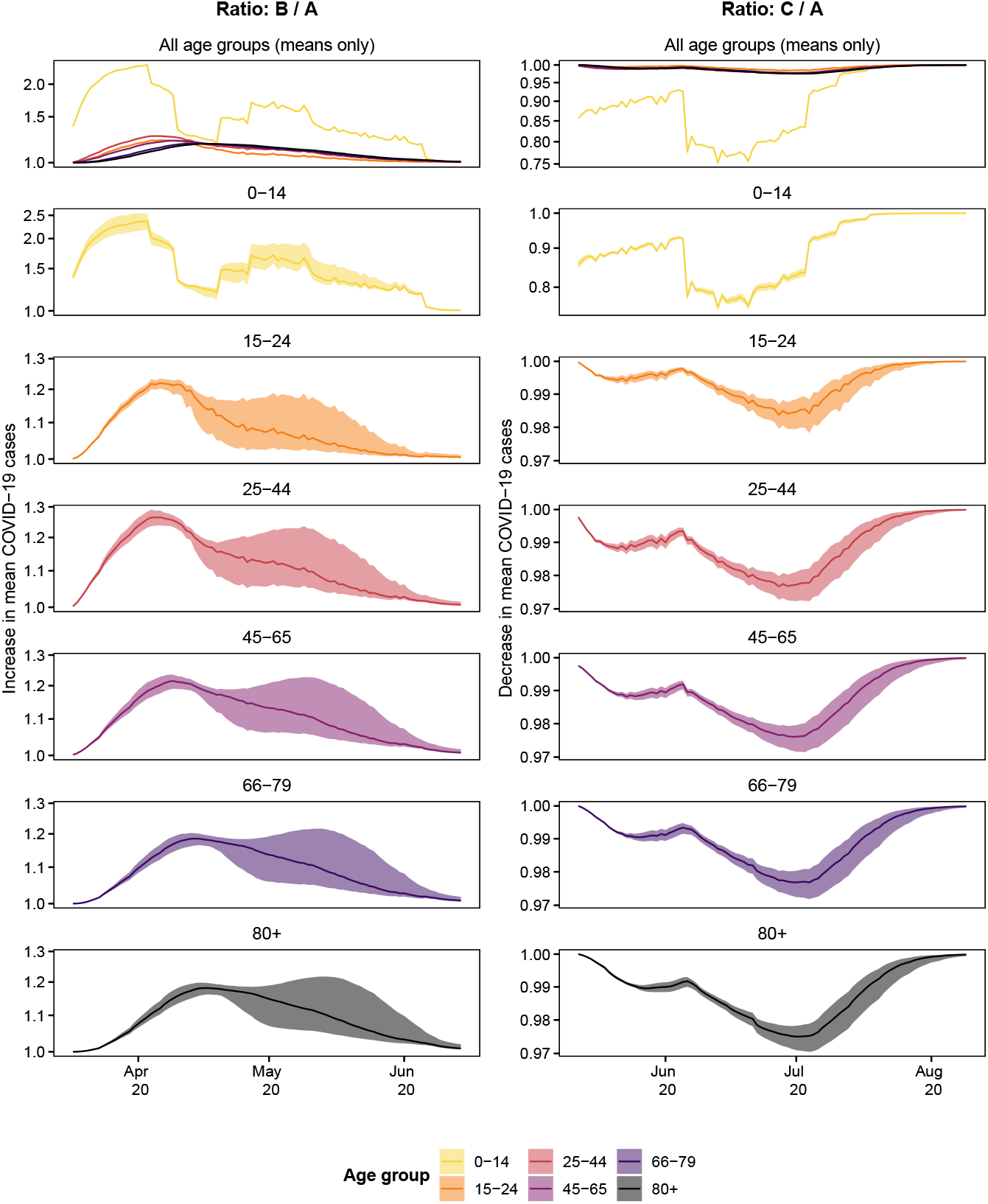
Showcased are the 10^th^, 50^th^, and 90^th^ percentiles of ratios of simulated path trajectories. The ratios of predicted mean cases for scenario B (schools open) divided by those predicted under scenario A (true measures, left) and for scenario C (schools closed) over the number of cases predicted in scenario A (right). The y-axis is log-transformed

**Figure 6:**
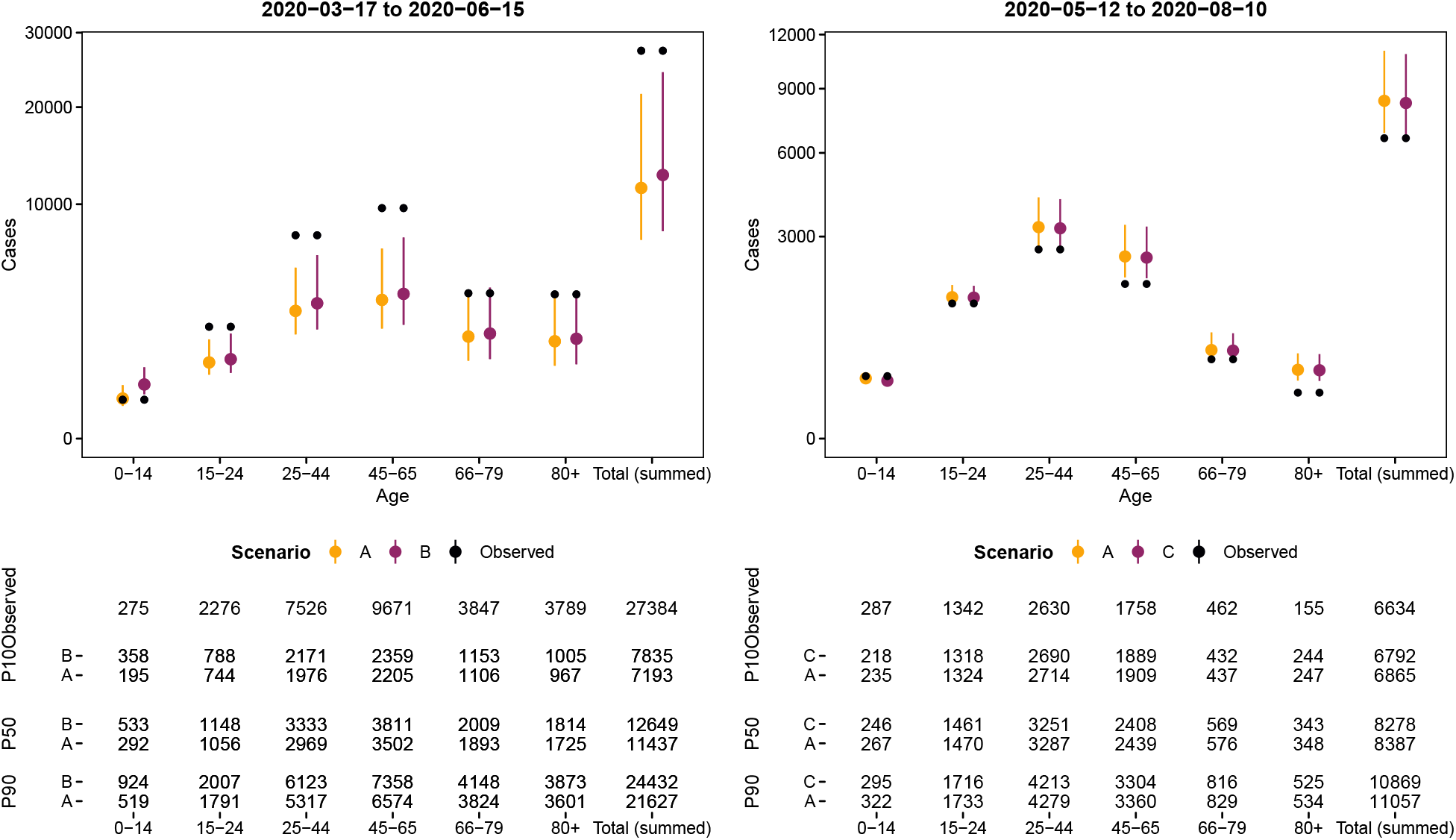
Comparison between simulated number of mean cases over 90 days from 17^th^ March 2020 for scenarios A (true measures) and Scenario B (schools open) and 90 days from 12^th^ May 2020 for scenarios A (true measures) and C (schools closed). Showcased are the 10^th^, 50^th^, and 90^th^ percentiles as well as the observed case counts in the period considered

The final epidemic size estimates (with uncertainty bounds) within each scenario are given in Figure 6 and are calculated by summing the predicted number of cases across the 90 day projection window. Table 3 shows the differences and ratios between expected cases of scenario A and scenarios B and C for the 90 day periods. Due to the time-sensitive nature of the policy questions being considered, the focus of this work was not calibration (the match of observations and predictions) but rather the differences in scenarios. We discuss the difficulties of forecasting in more detail in our discussion. We are most interested in the ratio between the predicted case counts: the percentage increase and decrease in cases is approximately ten per cent for both scenarios (the ratio is 1.11 for scenario B and 0.99 for scenario C). Most of the effect is found among the youngest group, which have 82 per cent more cases for scenario B and 8 per cent fewer cases for scenario C. The relative difference in expected cases between scenarios A and B suggests that case numbers would not have increased a lot of schools were left open and regarding scenario C, as expected; closing schools decreases cases.

**Table 3:** Comparisons of the number of cases in scenario A (true measures) with the number of cases in scenarios B (schools open) between 17^th^ March 2020 and 90 days and C (schools closed) between 12^th^ May 2020 and 90 days

| Age | B - A |  |  | B / A |  |  | C - A |  |  | C / A |  |  |
| --- | --- | --- | --- | --- | --- | --- | --- | --- | --- | --- | --- | --- |
|  | P<sub>10</sub> | P<sub>50</sub> | P<sub>90</sub> | P<sub>10</sub> | P<sub>50</sub> | P<sub>90</sub> | P<sub>10</sub> | P<sub>50</sub> | P<sub>90</sub> | P<sub>10</sub> | P<sub>50</sub> | P<sub>90</sub> |
| 0-14 | 162.9 | 240.0 | 404 | 1.76 | 1.82 | 1.87 | -27.5 | -20.85 | -16.98 | 0.91 | 0.92 | 0.93 |
| 15-24 | 45.1 | 89.9 | 218 | 1.06 | 1.09 | 1.12 | -16.6 | -9.38 | -5.98 | 0.99 | 0.99 | 1.00 |
| 25-44 | 192.9 | 362.3 | 821 | 1.10 | 1.12 | 1.15 | -64.4 | -36.03 | -23.15 | 0.98 | 0.99 | 0.99 |
| 45-65 | 153.3 | 311.5 | 773 | 1.07 | 1.09 | 1.12 | -55.2 | -30.08 | -18.96 | 0.98 | 0.99 | 0.99 |
| 66-79 | 48.9 | 113.8 | 323 | 1.04 | 1.06 | 1.08 | -13.5 | -6.92 | -4.05 | 0.98 | 0.99 | 0.99 |
| 80+ | 37.3 | 90.5 | 271 | 1.04 | 1.05 | 1.07 | -9.5 | -4.69 | -2.62 | 0.98 | 0.99 | 0.99 |
| Total (summed) | 641.7 | 1,207.1 | 2,820 | 1.09 | 1.11 | 1.13 | -186.0 | -107.83 | -71.69 | 0.98 | 0.99 | 0.99 |

## 4 Discussion

In our earlier work attempting to provide evidence-based information for policy makers, we found than an endemic-epidemic model (a two-component model for infectious disease) provided a good fit for data from Zurich, Switzerland [Bekker-Nielsen Dunbar et al., 2022]. The model had effects of day of the week, public holidays, testing rate, and age in the endemic components and the same effects as well as a centred linear time trend in the endemic component. This model suggested if there was no school closure, an increase in cases would be expected in the youngest age group (0–14) in April and later in other age groups, with the next age group expected to experience an increase in cases compared with the true school closure implemented being the parents of the first age group (25–44; parents). In this work, the main group of concern was the oldest age group (80+) and they were found to be consistently the lowest in terms of expected increase in cases compared with what was expected when schools were closed. Here we extended this work further motivated by the fact the usefulness of school closure to combat COVID-19 was not fully determined by end of the “first wave”. Schools in Switzerland re-opened after the summer of 2020 but at the time questions of whether to close them remained. In our work we were able to examine school closure at greater spatial and temporal scale than previously which is a strength of the approach. Other countries were observed to have different levels of school closure during the study period compared with Switzerland and so the “ideal” amount of closure remains to be determined. We note that school closures are a primary measure for disease control but other measures such as masked students or vaccinated students which seek to reduce within-class disease risk may be a better option later in the outbreak [Endo et al., 2021]. We remain cognisant that the purpose of school is not just educational and it is important to investigate the impact of this as the knock-on effects to children’s health of remote learning are expected to be a topic of interest for years to come. While the current work considers only the options of schools open or closed, the methodology used could also be used to examine use of masks in educational settings, provided evidence is available to inform the timevarying transmission weights and so is very versatile.

In the age group 80+ we find that incidence is completely explained by the epidemic component *ϕ* so the endemic component was not identifiable and diverged. It is not ideal that 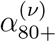 was restricted to be zero but the alternative approach of setting all *α*^(*ν*)^ values to be the same would also not have been ideal since the results imply they differ across age groups (Table 2). The analysis we present is ecological as we aggregated our indicators across the federation of Switzerland although they differ across regions. Ideally we would have liked to have done a spatio-temporal analysis across age groups but as we were interested in specific age groups rather than ten-year age bands, we had to choose to focus on age over age and space.

The uncertainty shown in the plot is the uncertainty of the predicted mean. Scenario C has more data to predict in the one-step prediction approach used to calculate this mean due to its prediction window starting later. We suspect fewer cases early on in the study period to be the cause of more uncertainty in the left panels of Figure 5. There is less uncertainty on the predictions in scenario C but we also observed fewer cases (the incidence was low in the summer). Due to the time-sensitive nature of the policy questions being considered, the focus of this work was not calibration (the match of observations and predictions) but rather the differences in scenarios. However, we note that there are greater discrepancies between the predicted number of cases and the true number of cases for the real scenario in the analysis of scenario B (Figure 6). Scenario B underestimates the number of cases while Scenario C sometimes overestimates the number while the total number of observed cases is within the predictions. The predicted means are made on the basis of the same model which was fit over the entire study period (Figure 1). The reason for predicting a 90 day window rather than the entire year is that we find it unlikely that a decision maker at a public health agency would not revisit a decision made within a 90 day period, so predicting cases until the end of the period the model is fit on strikes us as a less useful exercise. Additionally the biology of the disease changed due to a different variant of the disease becoming dominant (“delta”/B.1.617.2) in the ecological niche and this information is not included in our modelling work. Indeed, the model does not have change points and the large increase in cases at the end of the study period (which the model is fit to) might influence its ability to predict lower case counts.

We briefly summarise the comparison of results with the Zurich analysis [see Bekker-Nielsen Dunbar et al., 2022, for details]. By virtue of the shorter time frame of the earlier analysis, we are unable to note similarities and differences with scenario C as this information is not available at Zurich level. The model for Zurich contains more effects than the model used here. The Zurich model 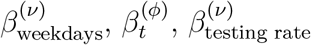 and 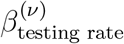. Notably, the Zurich analysis has additional time effects while our current model only contains a non-linear time trend in the form of the sine-cosine waves. While the models are different, the estimated discrete-time serial interval *û*_*d*_ is similar. Some of the building blocks used to construct the models are the same for the two studies: *p*_*s,t*_ and *d*_*s*_ are the same in the two studies. The relative increase (determined by calculating the ratio of predicted cases under scenario B and scenario A) takes values closer to 1 (no difference) for all age groups but the youngest. For Zurich the relative increase in these age groups is no more than five per cent, while it is slightly larger for the current work (ranging from 1.05 to 1.82 compared to 1.01 to 1.05). Many of the *P*_90_ values in Table 3 are a ten-fold increase with those found for Zurich. The ratio for the youngest age group (0–14) is much greater in the current work with no overlap with the values found in the previous work.

Finally we note that the existence of pharmaceutical countermeasures does not guarantee their use. Vaccines are recommended to prevent disease, disability, and death in children [World Health Organization, 2022]. However, with novel vaccines for pandemic control, children may be included in secondary but not primary trials and so may not be included in immunisation programmes as soon as a prophylaxis is tested safe and made available to the population. For this reason, we believe gaining an understanding of the impact of school closures in the absence of vaccine to still be an interesting and relevant area of research.

## Data Availability Statement

The protocol for this study can be accessed at https://osf.io/fgrdy which includes description of where to source data used. The code used in this work can be accessed at https://gitlab.switch.ch/suspend/COVID-19-school-CH. The majority of the data used in this work is publicly available; descriptions and access options can be found in the study protocol [Bekker-Nielsen Dunbar et al., 2022].

## Data Availability

The protocol for this study can be accessed at https://osf.io/fgrdy which includes description of where to source data used.

## Acknowledgements

This work is funded by the Swiss National Science Foundation (https://data.snf.ch/covid-19/snsf/196247). The authors declare no conflicts of interest besides some being guardians of children and some being residents of Switzerland and so have an interest in determining the overall benefit of school closures.

