## Supporting information for "Endemic-epidemic modelling of school closure to prevent spread of COVID-19 in Switzerland"

### Supporting information for "Endemic-epidemic modelling for school closure to prevent spread of COVID-19 in Switzerland"

This document contains ancillary information which is relevant to the manuscript. It consists of additional figures, an outline of the model selection procedure used, and a sensitivity analysis of time-varying transmission weights used in the modelling approach.

#### 1 Additional figures

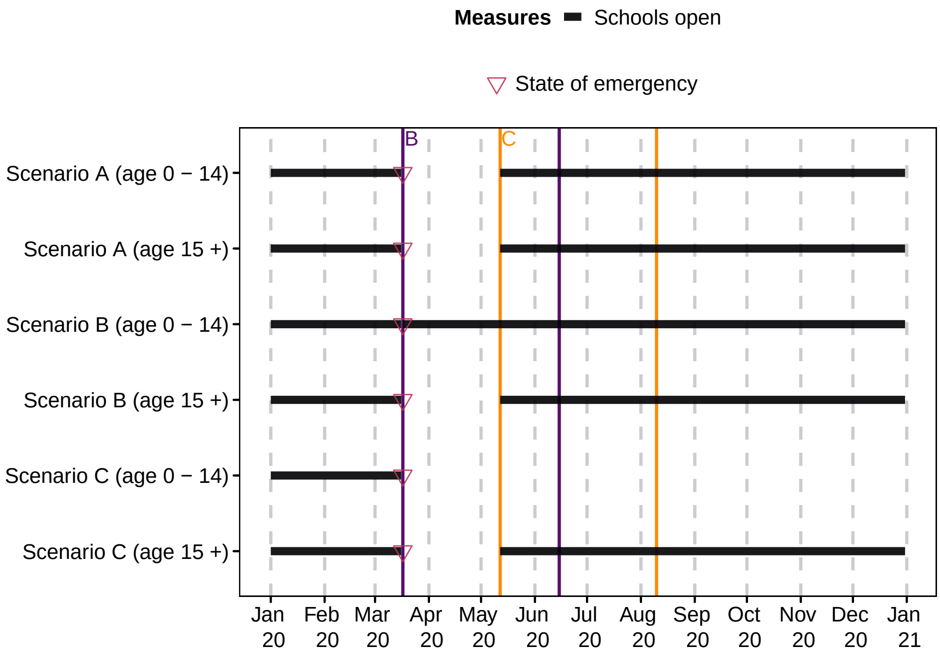


Figure S1: Illustration of scenarios. The state of emergency in Switzerland leading to "lockdown" was declared on $17^{th}$ March 2020 (vertical line between March and April 2020). The projection windows are shown with coloured vertical lines

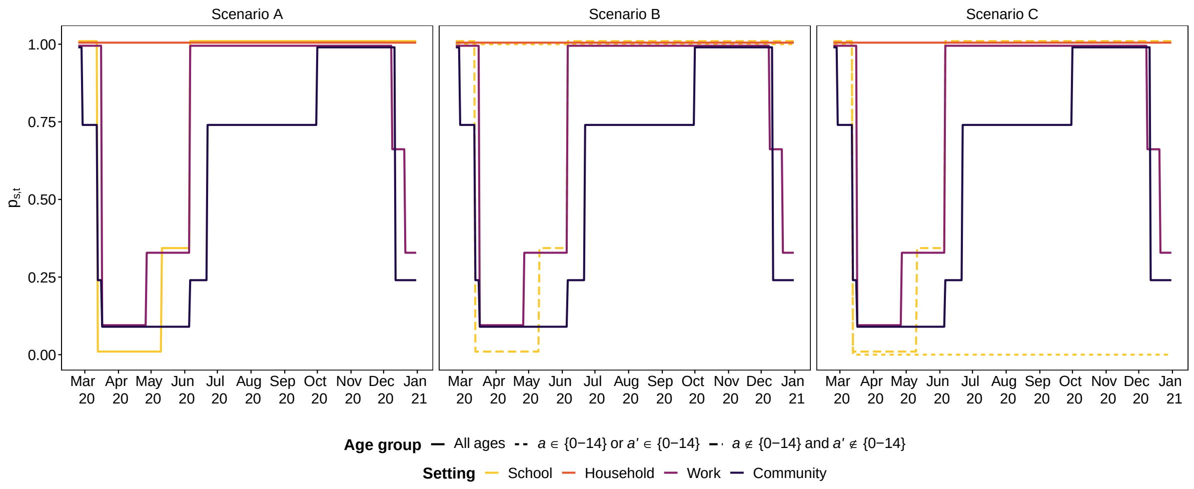


Figure S2: Policy information $p_{s,t,a,a'}$ for the scenarios considered. A slight jitter has been applied to assist in the visualisation

#### 2 Model selection procedure

The reference model is

$$Y_{at} \mid Y_{a,t-1}, \ldots, Y_{a,t-d_{\text{max}}} \sim \text{NegBin}(\lambda_{at}, \psi)$$

$$\lambda_{at} = \underbrace{\nu_{at} e_a}_{\text{endemic}} + \underbrace{\phi_{at} \sum_{a'} \sum_{d=1}^{d_{\text{max}}} u_d w_{a,a',t} Y_{a',t-d}}_{\text{epidemic}} \quad (1)$$

$$\log(\nu_{at}) = \alpha_a^{(\nu)} + \beta^{(\nu)}_{\text{public holiday}} x_t \quad (2)$$

$$\log(\phi_{at}) = \alpha_a^{(\phi)} + \beta^{(\phi)}_{\text{public holiday}} x_t \quad (3)$$

$$u_d \propto \frac{\kappa^{d-1}}{(d-1)!} \cdot \exp(-\kappa), \quad \kappa > 0, \quad d = 1, \ldots, d_{\text{max}}$$

This is denoted model 0 in our statistical analysis plan (can be found in our study protocol at https://osf.io/fgrdy). The reference model has fixed effects of age group, public holiday score, and school holiday score in both components. The model fit is shown in Figure S4. Overall, the predicted values (shaded area) seem to fit the observed data (points) quite well. During the first wave (here we consider cases before 1st June 2020 to be the first oscillation in the absence of a true definition of epidemic waves), the model overestimates the number of cases for all age groups except the youngest age group. This is seen in the shaded area being greater than the observed counts. We can also see that there might be some

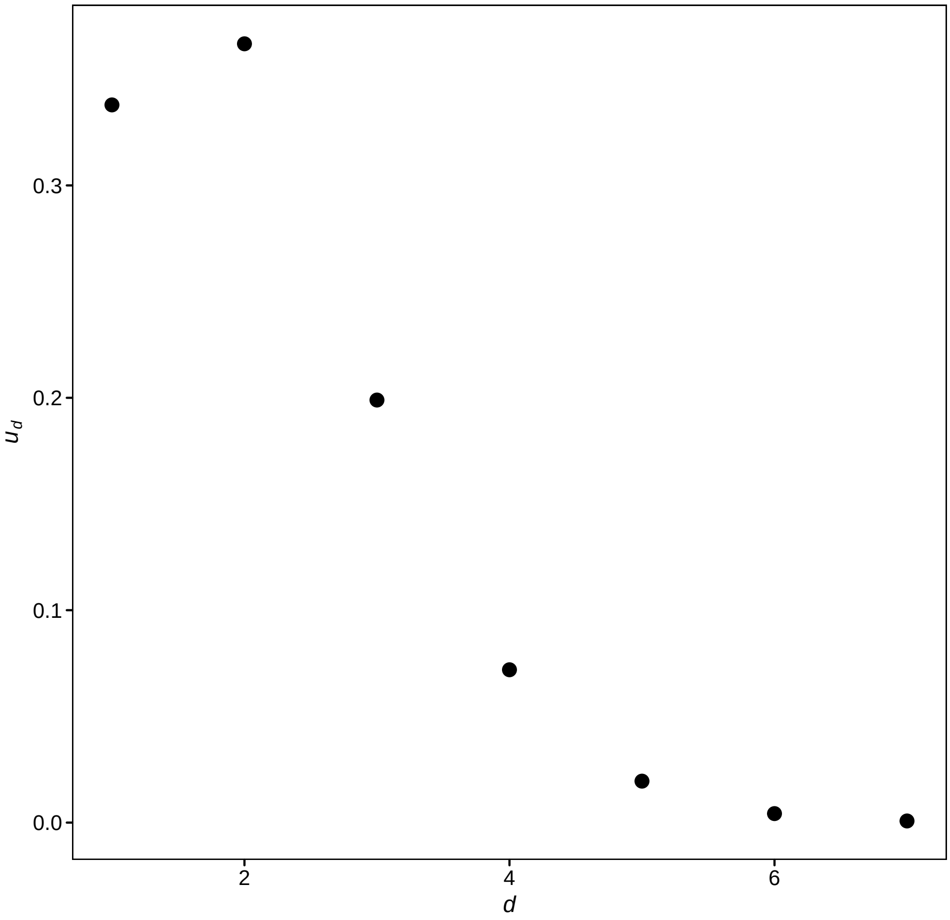


Figure S3: Estimated lag distribution $\hat{u_d}$ (below)

uncaptured weekday effects as the observed cases seem to oscillate more within a week than is captured by the model. According to the reference model, nearly all cases can be captured by the epidemic component $\phi$.

The model parameter estimates for the reference model are given in Table S1. Here we also experienced having a large standard error for the fixed effect of the oldest age group in the endemic component $\alpha^{(\nu)}_{80+}$. Dropping this coefficient from the model resolved the issue. This model has smaller values for $\alpha^{(\nu)}$ than the final model selected (estimates are shown in the main manuscript). The public holiday estimates are greater in this model. This implies some of the effect they capture here is explained by other effects when they are included in the model, such as those capturing the weekly fluctuations in cases. Overdispersion $\psi$ is also much larger than in the final model and the decreasing pattern found for $\alpha^{(\phi)}$ is not evid-

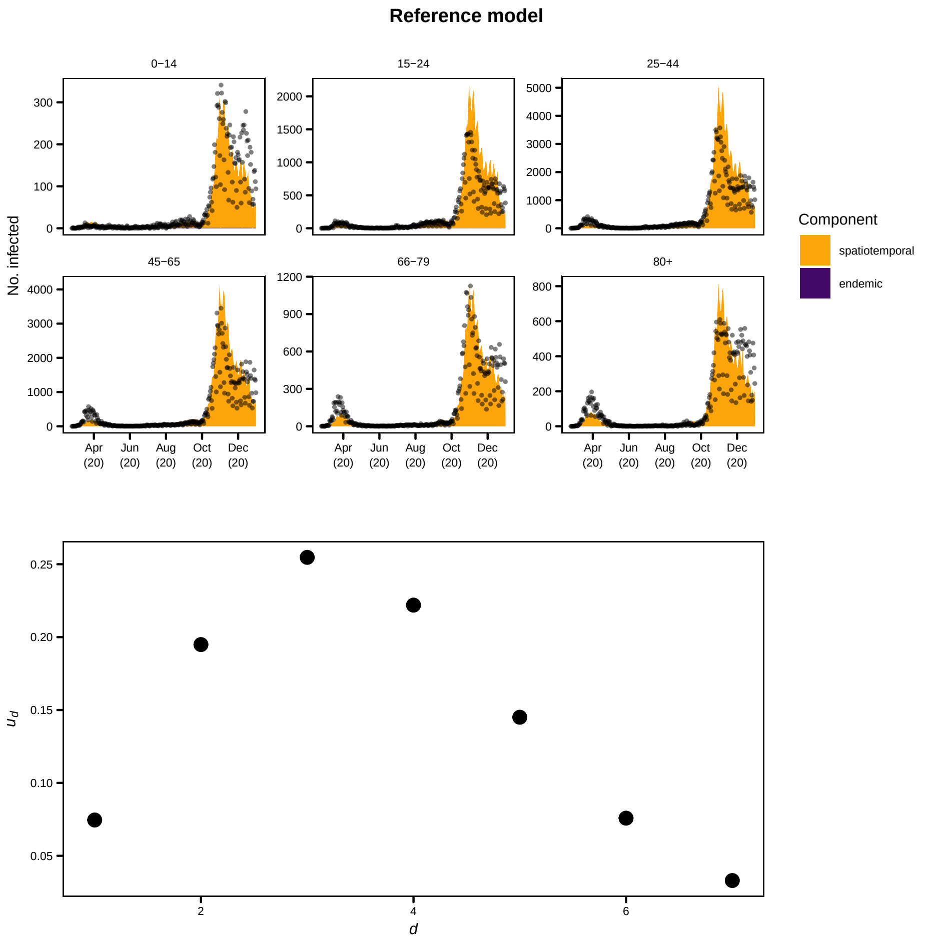


Figure S4: Reference model with age group specific effects as well as public holiday score in both components

ent for the reference model.

When constructing models in addition to the effects considered in the reference model, we considered covariates for sine-cosine waves (non-linear trend), temperature, testing rate, time trend (linear trend), weekday, and weekend. As certain covariates capture similar effects we did not fit models for all possible predictor combinations. Mutually exclusive effects within the same model component are weekday and weekend as well as temperature and non-linear trend represented through sine-cosine waves. The remaining covariates do not have any

#### Table S1: Reference model coefficients

| Endemic – $\log(\nu_{at})$ | | | Epidemic – $\log(\phi_{at})$ | | | Other parameters | | |
|---|---|---|---|---|---|---|---|---|
| Coefficient | Estimate | Std. Error | Coefficient | Estimate | Std. Error | Coefficient | Estimate | Std. Error |
| $\alpha^{(\nu)}_{0\text{-}14}$ | 2.002 | 0.178 | $\alpha^{(\phi)}_{0\text{-}14}$ | -3.607 | 0.045 | $\psi_{0-14}$ | 0.278 | 0.032 |
| $\alpha^{(\nu)}_{15\text{-}24}$ | 2.910 | 0.309 | $\alpha^{(\phi)}_{15\text{-}24}$ | -2.131 | 0.044 | $\psi_{15-24}$ | 0.318 | 0.028 |
| $\alpha^{(\nu)}_{25\text{-}44}$ | 1.907 | 0.522 | $\alpha^{(\phi)}_{25\text{-}44}$ | -1.983 | 0.040 | $\psi_{25-44}$ | 0.315 | 0.026 |
| $\alpha^{(\nu)}_{45\text{-}65}$ | 0.805 | 1.142 | $\alpha^{(\phi)}_{45\text{-}65}$ | -2.066 | 0.043 | $\psi_{45-65}$ | 0.389 | 0.032 |
| $\alpha^{(\nu)}_{66\text{-}79}$ | 1.273 | 0.627 | $\alpha^{(\phi)}_{66\text{-}79}$ | -1.802 | 0.043 | $\psi_{66-79}$ | 0.382 | 0.035 |
| | | | $\alpha^{(\phi)}_{80+}$ | -0.896 | 0.048 | $\psi_{80+}$ | 0.583 | 0.056 |
| $\beta^{(\nu)}_{\text{public holiday}}$ | -1.944 | 1.396 | $\beta^{(\phi)}_{\text{public holiday}}$ | -0.407 | 0.110 | | | |
| | | | | | | $\log\kappa$ | 0.961 | |

restrictions. Bayesian information criterion was used to determine goodness-of-fit of the models considered in this work. Table S2 shows the Bayesian information criterion values for the models considered. The selected model presented in the main manuscript is the model with option 14 for $\log(\nu_{at})$ and option 13 for $\log(\phi_{at})$ as described in our statistical analysis plan, which is available at https://osf.io/fgrdy..

#### Table S2: Models ranked by Bayesian information criterion (BIC)

| | Endemic *nu* | | | | | | Epidemic *phi* | | | | | | |
|---|---|---|---|---|---|---|---|---|---|---|---|---|---|
| Rank | Weekday | Weekend | Time trend | Testing rate | Temperature | Sine-cosine wave | Weekday | Weekend | Time trend | Testing rate | Temperature | Sine-cosine wave | BIC |
| 1 | | ✓ | | | | ✓ | ✓ | | | | | ✓ | 17,224 |
| 2 | ✓ | | | | | ✓ | ✓ | | | | | ✓ | 17,139 |
| 3 | | | ✓ | | | ✓ | ✓ | | | | | ✓ | 17,123 |
| 4 | | | | | | ✓ | ✓ | | | | | ✓ | 16,513 |
| 5 | | | ✓ | ✓ | | | ✓ | | | | | ✓ | 17,126 |
| 6 | | | ✓ | | | ✓ | ✓ | | | | ✓ | | 16,981 |
| 7 | | | ✓ | | | ✓ | ✓ | | | | | | 17,032 |
| 8 | | | ✓ | | | ✓ | ✓ | | | ✓ | | | 16,402 |
| 9 | | | ✓ | | | | ✓ | | | | | ✓ | 17,109 |
| 10 | | ✓ | | | ✓ | | ✓ | | | | | ✓ | 16,946 |
| 11 | | | ✓ | | ✓ | | ✓ | | | | | ✓ | 17,007 |
| 12 | | | | | ✓ | | ✓ | | | | | ✓ | 16,499 |
| 13 | | ✓ | | | | | ✓ | | | | | ✓ | 16,437 |
| 14 | ✓ | | | | ✓ | | ✓ | | | | | ✓ | 16,551 |
| 15 | | | | | | | ✓ | | | | | ✓ | 16,418 |
| 16 | ✓ | | | | | ✓ | | ✓ | | | | ✓ | 16,376 |
| 17 | ✓ | | | | | | ✓ | | | | | ✓ | 16,478 |
| 18 | | ✓ | | ✓ | | | ✓ | | | | | ✓ | 16,495 |
| 19 | | ✓ | | | | ✓ | | ✓ | | | | ✓ | 16,464 |
| 20 | | | ✓ | | | ✓ | | ✓ | | | | ✓ | 16,012 |
| 21 | | | | | | ✓ | | ✓ | | | | ✓ | 16,458 |
| 22 | | | ✓ | | ✓ | | | ✓ | | | | ✓ | 16,493 |
| 23 | | ✓ | | | | ✓ | ✓ | | | | ✓ | | 16,459 |
| 24 | ✓ | | | | | ✓ | ✓ | | | | ✓ | | 15,924 |
| 25 | | ✓ | | | | ✓ | ✓ | | | ✓ | | | 16,335 |
| 26 | | | ✓ | | | ✓ | | ✓ | | | | | 16,374 |

#### Table S2: Models ranked by Bayesian information criterion (BIC) *(continued)*

| | Endemic $nu$ | | | | | | Epidemic $phi$ | | | | | | |
|---|---|---|---|---|---|---|---|---|---|---|---|---|---|
| Rank | Weekday | Weekend | Time trend | Testing rate | Temperature | Sine-cosine wave | Weekday | Weekend | Time trend | Testing rate | Temperature | Sine-cosine wave | BIC |
| 27 | | | ✓ | | | ✓ | | ✓ | | | ✓ | | 16,341 |
| 28 | | | ✓ | | | ✓ | | ✓ | | ✓ | | | 16,012 |
| 29 | | | | | | ✓ | ✓ | | | | ✓ | | 15,871 |
| 30 | ✓ | | | | | ✓ | ✓ | | | ✓ | | | 15,852 |
| 31 | | ✓ | | | | ✓ | ✓ | | | | | | 15,821 |
| 32 | | | ✓ | ✓ | | | ✓ | | | | ✓ | | 15,590 |
| 33 | ✓ | | | | | ✓ | ✓ | | | | | | 16,591 |
| 34 | | | | | | ✓ | ✓ | | | ✓ | | | 16,599 |
| 35 | | | | | | ✓ | ✓ | | | | | | 16,582 |
| 36 | | | ✓ | ✓ | | | ✓ | | | ✓ | | | 16,121 |
| 37 | | | ✓ | ✓ | | | ✓ | | | | | | 16,561 |
| 38 | | ✓ | | | ✓ | | | ✓ | | | | ✓ | 16,577 |
| 39 | | | | | ✓ | | | ✓ | | | | ✓ | 16,566 |
| 40 | | ✓ | | | | | | ✓ | | | | ✓ | 16,035 |
| 41 | | | | | | | | ✓ | | | | ✓ | 16,451 |
| 42 | | | ✓ | | | | ✓ | | | | ✓ | | 16,465 |
| 43 | | ✓ | | | | ✓ | | ✓ | | ✓ | | | 16,456 |
| 44 | | | ✓ | | ✓ | | ✓ | | | | ✓ | | 16,115 |
| 45 | ✓ | | | | ✓ | | | ✓ | | | | ✓ | 16,010 |
| 46 | ✓ | | | | | | | ✓ | | | | ✓ | 16,008 |
| 47 | ✓ | | | | | ✓ | | ✓ | | ✓ | | | 15,988 |
| 48 | | | ✓ | ✓ | | | | ✓ | | | ✓ | | 15,804 |
| 49 | | ✓ | | | | ✓ | | ✓ | | | | | 17,033 |
| 50 | | | | | | ✓ | | ✓ | | ✓ | | | 17,062 |
| 51 | ✓ | | | | | ✓ | | ✓ | | | | | 17,028 |
| 52 | | | | | | ✓ | | ✓ | | | | | 16,818 |
| 53 | | | ✓ | | | | ✓ | | | | | | 17,037 |
| 54 | | | ✓ | | ✓ | | ✓ | | | | | | 16,715 |
| 55 | ✓ | | | | | ✓ | | ✓ | | | ✓ | | 17,045 |
| 56 | | ✓ | | | | ✓ | | ✓ | | | ✓ | | 16,933 |
| 57 | | | ✓ | ✓ | | | | ✓ | | | | | 16,928 |
| 58 | | | | | | ✓ | | ✓ | | | ✓ | | 16,875 |
| 59 | | | ✓ | | | | | ✓ | | | ✓ | | 16,897 |
| 60 | | | ✓ | | ✓ | | | ✓ | | | | | 16,824 |
| 61 | | | ✓ | | | | | ✓ | | | | | 16,496 |
| 62 | | ✓ | | | | ✓ | | | ✓ | ✓ | | | 16,336 |
| 63 | ✓ | | | | | ✓ | | | ✓ | | | ✓ | 16,503 |
| 64 | ✓ | | | | | ✓ | | | | | | ✓ | 16,744 |
| 65 | ✓ | | | | | ✓ | | | ✓ | ✓ | | | 17,222 |
| 66 | | ✓ | | | | ✓ | | | ✓ | | | ✓ | 17,116 |
| 67 | ✓ | | | | | ✓ | | | ✓ | | ✓ | | 17,100 |
| 68 | | | ✓ | | | | ✓ | | | ✓ | | | 17,298 |
| 69 | | | ✓ | | ✓ | | ✓ | | | ✓ | | | 17,252 |
| 70 | | | | | ✓ | | ✓ | | | ✓ | | | 17,284 |
| 71 | | ✓ | | | | ✓ | | | | | | ✓ | 17,255 |
| 72 | | ✓ | | | ✓ | | ✓ | | | ✓ | | | 16,351 |
| 73 | | | ✓ | | | ✓ | | | ✓ | | | ✓ | 17,099 |
| 74 | | | ✓ | | ✓ | | | | ✓ | ✓ | | | |
| 75 | ✓ | | | | ✓ | | ✓ | | | ✓ | | | 17,066 |
| 76 | | | ✓ | | ✓ | | | ✓ | | | ✓ | | 17,275 |
| 77 | | ✓ | | | ✓ | | ✓ | | | | ✓ | | 16,441 |
| 78 | | | | | ✓ | | ✓ | | | | | | 16,387 |
| 79 | ✓ | | | | | ✓ | | | ✓ | | | | 16,359 |
| 80 | | | ✓ | | | ✓ | | | | ✓ | | | 16,339 |
| 81 | | ✓ | | | ✓ | | ✓ | | | | | | 16,446 |
| 82 | | | | | ✓ | | ✓ | | | | ✓ | | 16,470 |
| 83 | | ✓ | | ✓ | | | ✓ | | | | ✓ | | 16,439 |
| 84 | | | | ✓ | | | ✓ | | | | ✓ | | 16,263 |
| 85 | | ✓ | | | | | ✓ | | | | ✓ | | 16,388 |
| 86 | | | ✓ | ✓ | | | | | | ✓ | | | 16,419 |
| 87 | | | | | | ✓ | | | ✓ | | | ✓ | 16,395 |
| 88 | | ✓ | | | | ✓ | | | | ✓ | | | 15,907 |
| 89 | | | ✓ | | | | | | ✓ | | | ✓ | 16,282 |
| 90 | | | ✓ | | | ✓ | | | ✓ | ✓ | | | 16,330 |
| 91 | ✓ | | | | ✓ | | ✓ | | | | ✓ | | 16,290 |
| 92 | | ✓ | | | | ✓ | | | ✓ | | ✓ | | 16,270 |

#### Table S2: Models ranked by Bayesian information criterion (BIC) *(continued)*

| | Endemic *nu* | | | | | | Epidemic *phi* | | | | | | |
|---|---|---|---|---|---|---|---|---|---|---|---|---|---|
| Rank | Weekday | Weekend | Time trend | Testing rate | Temperature | Sine-cosine wave | Weekday | Weekend | Time trend | Testing rate | Temperature | Sine-cosine wave | BIC |
| 93 | | | | | | | ✓ | | | | ✓ | | 15,858 |
| 94 | ✓ | | | | ✓ | | ✓ | | | | | | 15,818 |
| 95 | | | ✓ | | | ✓ | | | | | | | 15,789 |
| 96 | ✓ | | | ✓ | | | ✓ | | | | ✓ | | 15,597 |
| 97 | | | ✓ | | | ✓ | | | | | ✓ | | 16,573 |
| 98 | | | ✓ | ✓ | | | | | | | ✓ | | 16,586 |
| 99 | | | ✓ | ✓ | | | | | ✓ | | ✓ | | 16,569 |
| 100 | | | ✓ | | | ✓ | | | | | | ✓ | 16,433 |
| 101 | ✓ | | | | | ✓ | | | | ✓ | | | |
| 102 | | | | ✓ | | | ✓ | | | ✓ | | | 16,592 |
| 103 | | | ✓ | ✓ | | | | | ✓ | ✓ | | | |
| 104 | ✓ | | | | | | ✓ | | | | ✓ | | 16,802 |
| 105 | | ✓ | | ✓ | | | ✓ | | | ✓ | | | 16,423 |
| 106 | | | | | | ✓ | | | ✓ | ✓ | | | |
| 107 | | | ✓ | ✓ | | | | | | | | | 16,439 |
| 108 | | ✓ | | | | ✓ | | | | | ✓ | | 16,919 |
| 109 | | | ✓ | | | | | ✓ | | | | ✓ | 15,990 |
| 110 | | ✓ | | | | ✓ | | | | | | | 15,966 |
| 111 | ✓ | | | ✓ | | | ✓ | | | ✓ | | | 15,948 |
| 112 | | | | | ✓ | | | ✓ | | ✓ | | | 15,809 |
| 113 | | | ✓ | ✓ | | | | ✓ | | | | ✓ | 16,652 |
| 114 | | | ✓ | | | | | ✓ | | ✓ | | | 16,616 |
| 115 | | | | | | ✓ | | | | | ✓ | | 16,578 |
| 116 | | | ✓ | | | ✓ | | | ✓ | | ✓ | | 16,445 |
| 117 | | | | | | ✓ | | | | | | | |
| 118 | | ✓ | | | ✓ | | | ✓ | | ✓ | | | 16,797 |
| 119 | | ✓ | | | | | ✓ | | | ✓ | | | 16,789 |
| 120 | | | | | | ✓ | | | | ✓ | | | 16,389 |
| 121 | ✓ | | | ✓ | | | | | ✓ | | | ✓ | 16,650 |
| 122 | | | ✓ | | | | | | ✓ | ✓ | | | |
| 123 | | | | | | | ✓ | | | ✓ | | | 16,574 |
| 124 | | | | | ✓ | | | ✓ | | | | | 16,301 |
| 125 | | ✓ | | | ✓ | | | ✓ | | | | | 16,396 |
| 126 | | | | ✓ | | | ✓ | | | | | | 16,178 |
| 127 | | ✓ | | ✓ | | | ✓ | | | | | | 16,150 |
| 128 | | | | ✓ | | | | ✓ | | | ✓ | | 16,364 |
| 129 | | ✓ | | ✓ | | | | ✓ | | | ✓ | | 17,139 |
| 130 | | ✓ | | | | | ✓ | | | | | | 16,996 |
| 131 | ✓ | | | | ✓ | | | ✓ | | | | | 16,991 |
| 132 | | | | | ✓ | | | ✓ | | | ✓ | | 16,503 |
| 133 | ✓ | | | | | | ✓ | | | ✓ | | | 17,222 |
| 134 | ✓ | | | ✓ | | | | ✓ | | | ✓ | | 16,955 |
| 135 | | ✓ | | | | | | ✓ | | | ✓ | | 17,220 |
| 136 | ✓ | | | | ✓ | | | ✓ | | | ✓ | | 16,384 |
| 137 | | | | | | | ✓ | | | | | | 17,127 |
| 138 | ✓ | | | ✓ | | | | | | | | ✓ | 16,944 |
| 139 | ✓ | | | ✓ | | | | | ✓ | | ✓ | | 16,985 |
| 140 | | | ✓ | | ✓ | | | | ✓ | | | ✓ | 16,496 |
| 141 | ✓ | | | | | | | ✓ | | | ✓ | | 16,435 |
| 142 | | | | | | | | ✓ | | | ✓ | | 16,551 |
| 143 | ✓ | | | ✓ | | | ✓ | | | | | | 16,416 |
| 144 | ✓ | | | | | | ✓ | | | | | | 16,384 |
| 145 | | | ✓ | | ✓ | | | | | | ✓ | | 16,371 |
| 146 | | | | | | ✓ | | | ✓ | | | | 16,392 |
| 147 | | | ✓ | | ✓ | | | | | | | | 16,349 |
| 148 | | ✓ | | | ✓ | | | | ✓ | | | ✓ | 15,944 |
| 149 | | ✓ | | | | ✓ | | | ✓ | | | | 16,347 |
| 150 | | | ✓ | | | | | | | | ✓ | | 16,377 |
| 151 | ✓ | | | | ✓ | | | | ✓ | | | ✓ | 16,343 |
| 152 | | | ✓ | | ✓ | | | | | | | ✓ | 15,849 |
| 153 | | | ✓ | | | | | | | | | | 16,341 |
| 154 | | | | ✓ | | | | | ✓ | | | ✓ | 16,365 |
| 155 | | | ✓ | | | | | | | | | ✓ | 16,331 |
| 156 | | ✓ | | ✓ | | | | | ✓ | | | ✓ | 15,951 |
| 157 | | | | | ✓ | | | | ✓ | | | ✓ | 15,813 |
| 158 | ✓ | | | | | ✓ | | | | | ✓ | | 15,780 |

#### Table S2: Models ranked by Bayesian information criterion (BIC) *(continued)*

| | Endemic $nu$ | | | | | | Epidemic $phi$ | | | | | | |
|---|---|---|---|---|---|---|---|---|---|---|---|---|---|
| Rank | Weekday | Weekend | Time trend | Testing rate | Temperature | Sine-cosine wave | Weekday | Weekend | Time trend | Testing rate | Temperature | Sine-cosine wave | BIC |
| 159 | ✓ | | | | | ✓ | | | | | | | 15,750 |
| 160 | | | | ✓ | | | | ✓ | | | | | 15,586 |
| 161 | | ✓ | | ✓ | | | | ✓ | | | | | 16,488 |
| 162 | | ✓ | | | | | | ✓ | | ✓ | | | 16,488 |
| 163 | | | | | | | | ✓ | | ✓ | | | 16,472 |
| 164 | | ✓ | | | ✓ | | | | ✓ | ✓ | | | 16,069 |
| 165 | ✓ | | | ✓ | | | | ✓ | | | | | 16,460 |
| 166 | | ✓ | | | | | | | ✓ | ✓ | | | 16,472 |
| 167 | | ✓ | | | | | | ✓ | | | | | 16,460 |
| 168 | | ✓ | | | | | | | ✓ | | | ✓ | 15,978 |
| 169 | ✓ | | | | | | | ✓ | | ✓ | | | 16,470 |
| 170 | | | | | | | | ✓ | | | | | 16,474 |
| 171 | ✓ | | | ✓ | | | | ✓ | | ✓ | | | 16,909 |
| 172 | ✓ | | | | | | | ✓ | | | | | 16,330 |
| 173 | ✓ | | | | | | | | ✓ | | | ✓ | 16,045 |
| 174 | | ✓ | | ✓ | | | | | ✓ | | ✓ | | 16,015 |
| 175 | ✓ | | | | | | | | ✓ | ✓ | | | 16,020 |
| 176 | | | ✓ | | ✓ | | | | ✓ | | ✓ | | 15,805 |
| 177 | | | | | | ✓ | | | | | | ✓ | 16,834 |
| 178 | | | | | | | | | ✓ | | | ✓ | 16,853 |
| 179 | | | | | ✓ | | | | ✓ | ✓ | | | 16,825 |
| 180 | | | | | | | | | ✓ | ✓ | | | 16,793 |
| 181 | | ✓ | | ✓ | | | | | | | | ✓ | 16,761 |
| 182 | | | ✓ | ✓ | | | | | ✓ | | | ✓ | 16,483 |
| 183 | | ✓ | | | ✓ | | | | | | | ✓ | 16,611 |
| 184 | | | ✓ | ✓ | | | | | | | | ✓ | 16,385 |
| 185 | | | | | | ✓ | | | ✓ | | ✓ | | 16,816 |
| 186 | ✓ | | | | ✓ | | | | | | | ✓ | 16,801 |
| 187 | | ✓ | | | | | | | | | | ✓ | 16,760 |
| 188 | ✓ | | | | | | | | | | | ✓ | 16,625 |
| 189 | ✓ | | | ✓ | | | | | ✓ | | | | 16,679 |
| 190 | | | ✓ | | | ✓ | | | ✓ | | | | 16,237 |
| 191 | | ✓ | | | ✓ | | | | ✓ | | ✓ | | 16,365 |
| 192 | | | | ✓ | | | | | ✓ | | ✓ | | 16,437 |
| 193 | | ✓ | | ✓ | | | | | ✓ | ✓ | | | 16,810 |
| 194 | | | ✓ | | | | | | ✓ | | ✓ | | 16,703 |
| 195 | ✓ | | | ✓ | | | | | ✓ | ✓ | | | 16,701 |
| 196 | | | | | ✓ | | | | | | | ✓ | 16,520 |
| 197 | ✓ | | | | ✓ | | | | ✓ | | ✓ | | 16,935 |
| 198 | | | ✓ | ✓ | | | | ✓ | | ✓ | | | 16,482 |
| 199 | | | | | | | | | | | | ✓ | 16,666 |
| 200 | | | | | ✓ | | | | ✓ | | ✓ | | 16,677 |
| 201 | | | ✓ | | | | | | ✓ | | | | 16,800 |
| 202 | | | ✓ | | ✓ | | | | ✓ | | | | 16,686 |
| 203 | | ✓ | | | | | | | ✓ | | ✓ | | 16,674 |
| 204 | | | | | | | | | ✓ | | ✓ | | 16,509 |
| 205 | ✓ | | | | | | | | ✓ | | ✓ | | 16,642 |
| 206 | ✓ | | | | ✓ | | | | ✓ | | | | 16,160 |
| 207 | | ✓ | | | ✓ | | | | ✓ | | | | 16,284 |
| 208 | | ✓ | | | ✓ | | | ✓ | | | ✓ | | 16,386 |
| 209 | | | ✓ | | ✓ | | | ✓ | | ✓ | | | 15,658 |
| 210 | | | | | ✓ | | | | ✓ | | | | 15,679 |
| 211 | | | ✓ | ✓ | | | | | ✓ | | | | 15,647 |
| 212 | | | | ✓ | | | | | | | | ✓ | 15,615 |
| 213 | ✓ | | | | ✓ | | | | | | ✓ | | |
| 214 | ✓ | | | | ✓ | | | | | | | | |
| 215 | ✓ | | | ✓ | | | | | | | ✓ | | 15,684 |
| 216 | ✓ | | | ✓ | | | | | | | | | 15,530 |
| 217 | | ✓ | | | ✓ | | | | | | ✓ | | 15,637 |
| 218 | | ✓ | | | | | | | | | ✓ | | 15,649 |
| 219 | ✓ | | | | | | | | | | ✓ | | 15,618 |
| 220 | | ✓ | | | ✓ | | | | | | | | 15,622 |
| 221 | | ✓ | | | | | | | ✓ | | | | 15,461 |
| 222 | | ✓ | | ✓ | | | | | | | | | 15,408 |
| 223 | | | | | | | | | ✓ | | | | 15,389 |
| 224 | | | | ✓ | | | | | ✓ | | | | 15,440 |

Table S2: Models ranked by Bayesian information criterion (BIC) *(continued)*

| | Endemic $nu$ | | | | | | Epidemic $phi$ | | | | | | |
|---|---|---|---|---|---|---|---|---|---|---|---|---|---|
| Rank | Weekday | Weekend | Time trend | Testing rate | Temperature | Sine-cosine wave | Weekday | Weekend | Time trend | Testing rate | Temperature | Sine-cosine wave | BIC |
| 225 | | ✓ | | ✓ | | | | | ✓ | | | | 15,944 |
| 226 | ✓ | | | | | | | | ✓ | | | | 15,962 |
| 227 | | ✓ | | | ✓ | | | | | ✓ | | | 15,938 |
| 228 | | | | | ✓ | | | | | ✓ | | | 16,418 |
| 229 | | ✓ | | | | | | | | ✓ | | | |
| 230 | | | | | ✓ | | | | | | | | |
| 231 | ✓ | | | | | | | | | ✓ | | | |
| 232 | | ✓ | | | | | | | | | | | 16,432 |
| 233 | | | | ✓ | | | | | | | | | 15,935 |
| 234 | | | | | ✓ | | | | | | ✓ | | 15,953 |
| 235 | ✓ | | | | | | | | | | | | 15,926 |
| 236 | | | | | | | | | | | ✓ | | 15,743 |
| 237 | | ✓ | | ✓ | | | | | | | ✓ | | 15,738 |
| 238 | | | | ✓ | | | | | | | ✓ | | 15,668 |
| 239 | | | | | | | | | | ✓ | | | 15,687 |
| 240 | | | | | | | | | | | | | 15,719 |
| 241 | | | | ✓ | | | | | | ✓ | | | 16,642 |
| 242 | | ✓ | | ✓ | | | | | | ✓ | | | 16,607 |
| 243 | | | ✓ | | ✓ | | | | | ✓ | | | 16,582 |
| 244 | ✓ | | | ✓ | | | | | | ✓ | | | 16,361 |
| 245 | | | ✓ | | | | | | | ✓ | | | 16,518 |
| 246 | ✓ | | | | ✓ | | | | | ✓ | | | 16,445 |
| 247 | | | | ✓ | | | | ✓ | | ✓ | | | 16,523 |
| 248 | | ✓ | | ✓ | | | | ✓ | | ✓ | | | 16,667 |
| 249 | ✓ | | | | ✓ | | | ✓ | | ✓ | | | 16,533 |
| 250 | | | | ✓ | | | | | ✓ | ✓ | | | 16,507 |
| 251 | ✓ | | | | ✓ | | | | ✓ | ✓ | | | 16,500 |
| 252 | | | | ✓ | | | ✓ | | | | | ✓ | 16,486 |
| 253 | ✓ | | | ✓ | | | ✓ | | | | | ✓ | 16,354 |
| 254 | | | | ✓ | | | | ✓ | | | | ✓ | 16,152 |
| 255 | ✓ | | | ✓ | | | | ✓ | | | | ✓ | 16,228 |
| 256 | | ✓ | | ✓ | | | | ✓ | | | | ✓ | 16,295 |

#### 3 Sensitivity analysis of contact matrices – model fit

The modelling approach used is dependent on the transmission weights $w_{a,a',t}$ used to inform the model. For this reason, we elect to do a sensitivity analysis. In particular, we consider it unrealistic that school contacts decrease and other contacts would be expected to remain the same so we conducted a sensitivity analysis of the situation where other contacts change to reflect this. We examine the robustness of the time-varying contact matrix $w_{a,a',t}$ through various alternatives for its construction. The total average contacts at time $t$ are given by the weighted sum

$$w_{a,a',t} = \sum_s \gamma_{s,t} \cdot c_{a,a',s} = \sum_s d_s \cdot p_{s,t} \cdot h_{s,t} \cdot c_{a,a',s} \tag{4}$$

where $s$ denotes setting the contact occurred in, $d_s$ are the disease-specific weights, $p_{s,t}$ is policy, and $h_{s,t}$ is adjustments for school holidays. The setting-specific contacts $c_{a,a',s}$ are the same as the ones displayed in Figure 2 in the main manuscript. The policy indicators $p_{s,t}$ used in this sensitivity analysis are the ones shown in Figure 3 of the main manuscript. We now consider the situation where $d_s$ are allowed to vary by time as well as setting: $d_{s,t}$. Previously we considered $d_s$ to take the values 11.41 for school setting, 8.07 for work setting, 4.11 for household, and 2.79 for other setting for all time points $t$. We conduct two sensitivity analyses to reflect that reductions in school contacts are unlikely to exist in a vacuum and there is likely to be some symbiosis with work contacts. Namely, when children are home from school guardians are more likely to also be home to engage in childcare. The two options for sensitivity analysis are:

Option 1: The household weight $d_s|_{\text{household}}$ reflects the school closure policy and is given by $d_s|_{\text{household}} = 2 - p_{s,t}|_{\text{school}}$

Option 2: The household weight $d_s|_{\text{household}}$ is adjusted by any amount which reduces contacts on school holidays (the adjustment is now through $h_{s,t}$)

#### 3.1 Option 1: Household contacts depend on school closures

The first option adjusts the household indicator such that it increases when the school indicator decreases. The rationale behind this is that when schools are closed, there are fewer school contacts but more household contacts so we considered that households should “mirror” schools to reflect expected increases in contacts due to school closures. The downside of this adjustment is that it does not take into account the school holiday score $h_{s,t}$ which means that if schools are generally open but it is a holiday in most cantons, household contacts are not increased (this will be addressed in Option 2). The implications of this adjustment on all involved components can be seen in Figure S5.

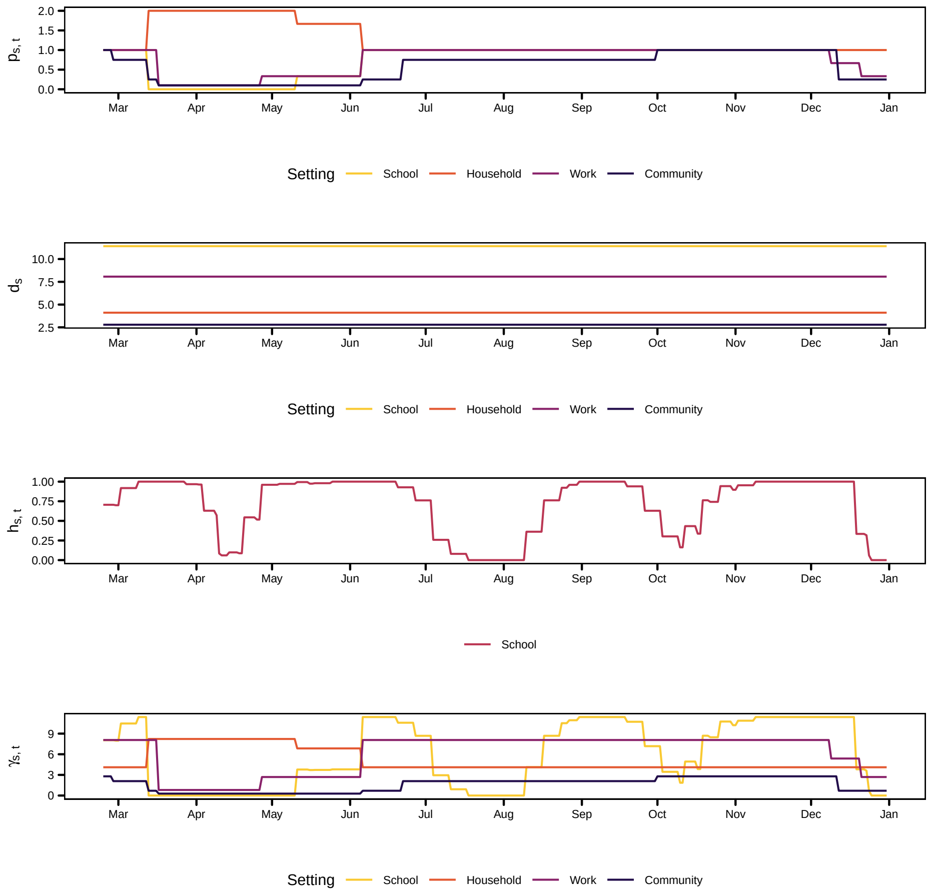


Figure S5: Time series of policy indicators $p_{s,t}$, disease weights $d_s$, and non-pandemic school holiday score $h_{s,t}$. The lower panel shows the product of all of the time series $\gamma_{s,t}$ which will serve as a multiplication factor for the setting specific contact matrix $c_{a,a',t}$

The fit and lag distribution of the model with contact matrices constructed under Option 1 is shown in Figure S6 and the corresponding parameters can be found in Table S3. Looking at the models fitted with the contact matrices constructed as explained under Option 1, the best model according to Bayesian information criterion has model covariates age, public holidays, weekend, and sine-cosine wave in the endemic component and age, public holidays, weekday, and sine-cosine wave in the epidemic component. The fit and lag distribution are shown in Figure S6.

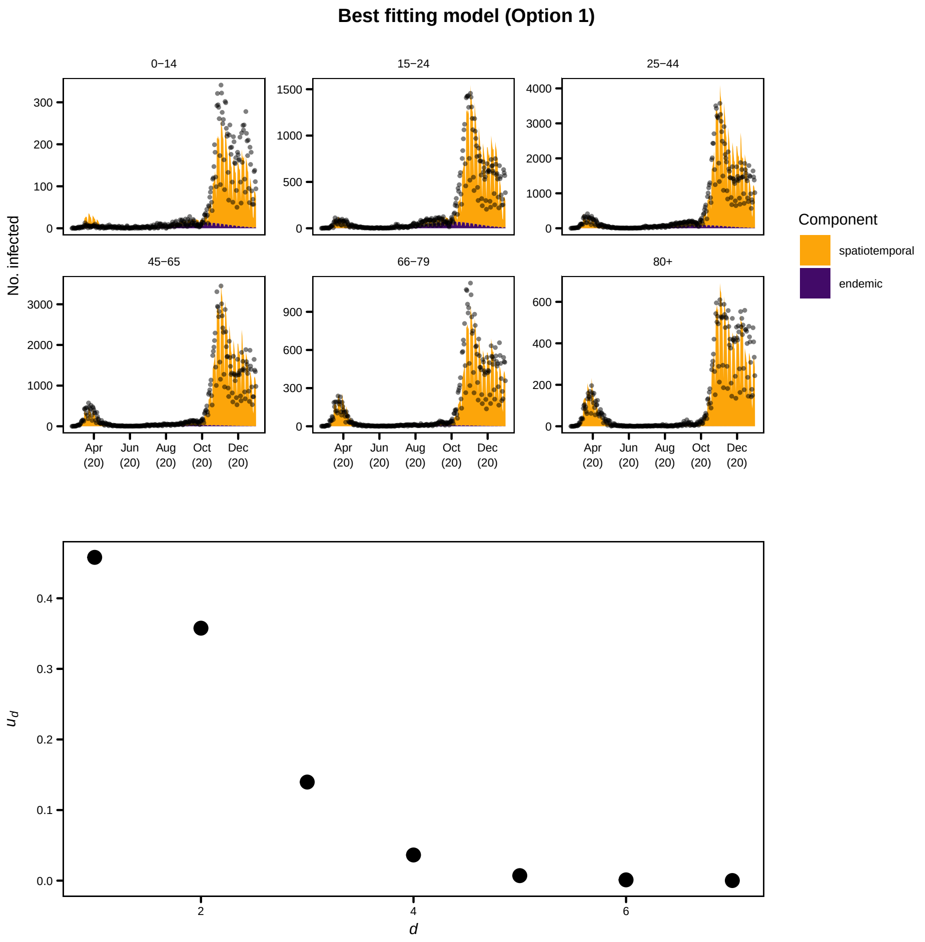


Figure S6: Model fit and lag distribution for the best model with weights as constructed under Option 1. The endemic component was fitted with effects for age, public holidays, weekend, and sine-cosine wave whereas the epidemic mean is fitted with effects for age, public holidays, weekday, and sine-cosine wave

When it comes to Option 1, we can see from the fit shown in Figure S6 that the number of cases in the youngest age group is clearly overestimated in the first wave (seen around April where the shaded area is above the points) and underestimated in the second wave (where the shaded area is below the points). The plot also shows that the contribution of the endemic mean $\hat{\nu}_{at}$ contributes predominantly in the younger age groups whereas for the older age groups almost all of the cases are attributed to the epidemic component $\hat{\phi}_{at}$. Furthermore, the estimated lag distribution $\hat{u}_d$ indicates that the maximum lag peaks at 1 which is not what we

Table S3: Coefficients of the best model using BIC under Option 1 and Scenario A.

| Endemic – $\log(\nu_{at})$ | | | Epidemic – $\log(\phi_{at})$ | | | Other parameters | | |
|---|---|---|---|---|---|---|---|---|
| Coefficient | Estimate | Std. Error | Coefficient | Estimate | Std. Error | Coefficient | Estimate | Std. Error |
| $\alpha^{(\nu)}_{\text{0-14}}$ | 2.617 | 0.195 | $\alpha^{(\phi)}_{\text{0-14}}$ | -3.995 | 0.054 | $\psi_{0-14}$ | 0.256 | 0.032 |
| $\alpha^{(\nu)}_{\text{15-24}}$ | 4.601 | 0.175 | $\alpha^{(\phi)}_{\text{15-24}}$ | -2.673 | 0.038 | $\psi_{15-24}$ | 0.116 | 0.012 |
| $\alpha^{(\nu)}_{\text{25-44}}$ | 3.878 | 0.177 | $\alpha^{(\phi)}_{\text{25-44}}$ | -2.371 | 0.026 | $\psi_{25-44}$ | 0.059 | 0.006 |
| $\alpha^{(\nu)}_{\text{45-65}}$ | 2.750 | 0.230 | $\alpha^{(\phi)}_{\text{45-65}}$ | -2.392 | 0.023 | $\psi_{45-65}$ | 0.051 | 0.005 |
| $\alpha^{(\nu)}_{\text{66-79}}$ | 2.302 | 0.259 | $\alpha^{(\phi)}_{\text{66-79}}$ | -2.134 | 0.028 | $\psi_{66-79}$ | 0.079 | 0.010 |
| | | | $\alpha^{(\phi)}_{\text{80+}}$ | -1.195 | 0.019 | $\psi_{80+}$ | 0.049 | 0.008 |
| | | | $\beta^{(\phi)}_{\text{day of the week Tuesday}}$ | 0.313 | 0.021 | | | |
| | | | $\beta^{(\phi)}_{\text{day of the week Wednesday}}$ | 0.067 | 0.022 | | | |
| | | | $\beta^{(\phi)}_{\text{day of the week Thursday}}$ | -0.053 | 0.022 | | | |
| | | | $\beta^{(\phi)}_{\text{day of the week Friday}}$ | -0.024 | 0.022 | | | |
| | | | $\beta^{(\phi)}_{\text{day of the week Saturday}}$ | -0.405 | 0.023 | | | |
| | | | $\beta^{(\phi)}_{\text{day of the week Sunday}}$ | -0.629 | 0.024 | | | |
| $\beta^{(\nu)}_{\text{weekend}}$ | -0.863 | 0.090 | | | | | | |
| $\beta^{(\nu)}_{\text{public holiday}}$ | -0.748 | 0.411 | $\beta^{(\phi)}_{\text{public holiday}}$ | -0.208 | 0.062 | | | |
| $\gamma^{(\nu)}$ | 1.927 | 0.185 | $\gamma^{(\phi)}$ | 0.557 | 0.019 | | | |
| $\delta^{(\nu)}$ | -2.233 | 0.027 | $\delta^{(\phi)}$ | 1.849 | 0.011 | | | |
| | | | | | | $\log \kappa$ | -0.247 | |

would expect. Compared with the original model in the main manuscript there is now a greater effect of $\beta^{(\nu)}_{\text{day of the week Friday}}$.

#### 3.2 Option 2: Weight shift from school to household during school holidays

The second option adjusts the household weight based on the reduction of the school weight through the holiday score. The reasoning behind this is that, contrary to the approach in Option 1, the contribution of household contacts expected to increase during school holidays. This means that the school weight decreases with the holiday score (as can be seen in Equation (4)) and the difference from the original school weight to the holiday score-adjusted is added to the household weight. The corresponding equation is:

$$d_{s,t}|_{\text{household}} = d_{s,t}|_{\text{household}} + c \cdot (d_{s,t}|_{\text{school}} - d_{s,t}|_{\text{school}} \cdot h_{s,t}) \qquad (5)$$

where $c$ denotes the fraction of reduced weight in the school setting that is added to the household setting. For this analysis we set $c = 0.5$ as we assume that the number of contacts is lower when individuals stay at home instead of going to school. The effect is shown in Figure S7.

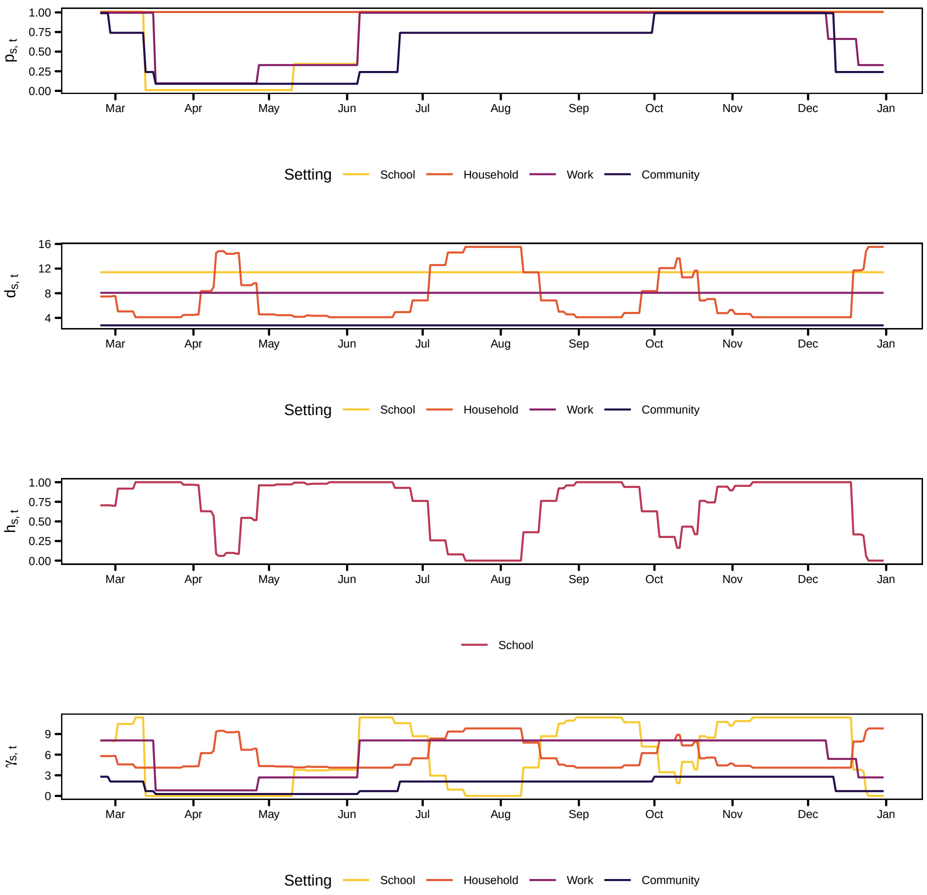


Figure S7: Time series of policy indicators $p_{s,t}$, disease weights $d_s$, and non-pandemic school holiday score $h_{s,t}$. The lower panel shows the product of all of the time series $\gamma_{s,t}$ which will serve as a multiplication factor for the setting specific contact matrix $c_{a,a',t}$

The corresponding plot and table for Option 2 can be found in Figure S8 and Table S4.

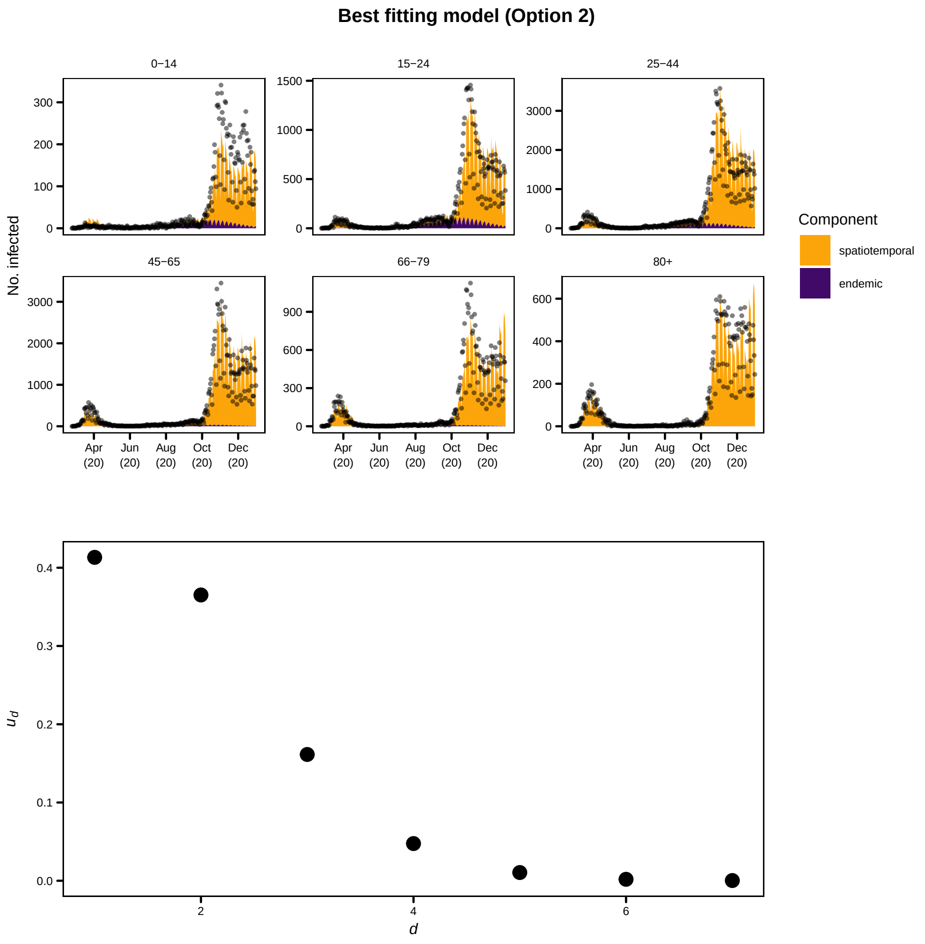


Figure S8: Model fit and lag distribution for the best model with transmission weights $w_{a,a',t}$ as constructed under Option 1. The endemic mean is fitted with effects for age, public holidays, weekday, and sine-cosine wave whereas the epidemic mean is fitted with effects for age, public holidays, weekday, and sine-cosine wave

The model fit shown in Figure S8 is similar to that seen for Option 1. However, the reduction in cases in November and December is not captured as well. Once again, the peak of the estimated lag distribution is at 1. Now the model contains weekday effects in the endemic component (Table S4). Thursday seems a particularly important day for case reporting as it has opposite effects in the two components. The overdispersion parameter $\nu$ takes low values but still shows some

Table S4: Coefficients of the best model using BIC under Option 2 and Scenario A

| Endemic – $\log(\nu_{at})$ | | | Epidemic – $\log(\phi_{at})$ | | | Other parameters | | |
|---|---|---|---|---|---|---|---|---|
| Coefficient | Estimate | Std. Error | Coefficient | Estimate | Std. Error | Coefficient | Estimate | Std. Error |
| $\alpha^{(\nu)}_{\text{0-14}}$ | 2.520 | 0.197 | $\alpha^{(\phi)}_{\text{0-14}}$ | -3.950 | 0.057 | $\psi_{0-14}$ | 0.231 | 0.031 |
| $\alpha^{(\nu)}_{\text{15-24}}$ | 4.574 | 0.152 | $\alpha^{(\phi)}_{\text{15-24}}$ | -2.619 | 0.036 | $\psi_{15-24}$ | 0.090 | 0.010 |
| $\alpha^{(\nu)}_{\text{25-44}}$ | 3.797 | 0.156 | $\alpha^{(\phi)}_{\text{25-44}}$ | -2.302 | 0.024 | $\psi_{25-44}$ | 0.054 | 0.006 |
| $\alpha^{(\nu)}_{\text{45-65}}$ | 2.512 | 0.250 | $\alpha^{(\phi)}_{\text{45-65}}$ | -2.315 | 0.023 | $\psi_{45-65}$ | 0.062 | 0.006 |
| $\alpha^{(\nu)}_{\text{66-79}}$ | 2.017 | 0.301 | $\alpha^{(\phi)}_{\text{66-79}}$ | -2.058 | 0.029 | $\psi_{66-79}$ | 0.099 | 0.012 |
| | | | $\alpha^{(\phi)}_{\text{80+}}$ | -1.129 | 0.022 | $\psi_{80+}$ | 0.080 | 0.013 |
| $\beta^{(\nu)}_{\text{day of the week Tuesday}}$ | 0.128 | 0.099 | $\beta^{(\phi)}_{\text{day of the week Tuesday}}$ | 0.355 | 0.025 | | | |
| $\beta^{(\nu)}_{\text{day of the week Wednesday}}$ | 0.362 | 0.087 | $\beta^{(\phi)}_{\text{day of the week Wednesday}}$ | 0.074 | 0.025 | | | |
| $\beta^{(\nu)}_{\text{day of the week Thursday}}$ | 0.410 | 0.084 | $\beta^{(\phi)}_{\text{day of the week Thursday}}$ | -0.066 | 0.025 | | | |
| $\beta^{(\nu)}_{\text{day of the week Friday}}$ | 0.255 | 0.094 | $\beta^{(\phi)}_{\text{day of the week Friday}}$ | 0.034 | 0.026 | | | |
| $\beta^{(\nu)}_{\text{day of the week Saturday}}$ | -0.322 | 0.103 | $\beta^{(\phi)}_{\text{day of the week Saturday}}$ | -0.454 | 0.026 | | | |
| $\beta^{(\nu)}_{\text{day of the week Sunday}}$ | -0.791 | 0.111 | $\beta^{(\phi)}_{\text{day of the week Sunday}}$ | -0.665 | 0.026 | | | |
| $\beta^{(\nu)}_{\text{public holiday}}$ | -0.167 | 0.316 | $\beta^{(\phi)}_{\text{public holiday}}$ | -0.706 | 0.071 | | | |
| $\gamma^{(\nu)}$ | 1.994 | 0.160 | $\gamma^{(\phi)}$ | 0.683 | 0.018 | | | |
| $\delta^{(\nu)}$ | -2.481 | 0.026 | $\delta^{(\phi)}$ | 1.515 | 0.013 | | | |
| | | | | | | $\log \kappa$ | -0.124 | |

age-dependence. The fixed effects of age group $\alpha^{(\phi)}$ are once again showing a decreasing pattern.

#### 3.3 Discussion

Our goodness-of-fit criterion for model selection–Bayesian information criterion (see previous section)– selects the same model as the main manuscript for Option 1 but a different model is selected for Option 2. This implies our results are robust to the shift in household contacts reflecting policy but not for that reflecting school holiday. Weekly fluctuations remain an important effect to capture in our models for daily case counts.

### 4 Sensitivity analysis of contact matrices – final size estimates

Considering the options outlined in the previous section, we examine the effect these changes to $\gamma_{s,t}$ has on our predicted case counts. The previous section considered only those results which are relevant for the model selection procedure, i.e. the model fit under scenario A and goodness-of-fit. Now we evaluate the predictions under the two alternative scenarios B and C. Recall, the two options for sensitivity analysis are:

Option 1: The household weight $d_s|_{\text{household}}$ reflects the school closure policy and is given by $d_s|_{\text{household}} = 2 - p_{s,t}|_{\text{school}}$ ("Household contacts depend on school closures")

Option 2: The household weight $d_s|_{\text{household}}$ is adjusted by any amount which reduces contacts on school holidays (the adjustment is now through $h_{s,t}$) ("Weight shift from school to household during school holidays")

#### 4.1 Scenario B

We now present the effect these changes have on the ratios of the predicted number of counts between Scenario A and Scenario B (Figure S9). We also compare the final size between scenarios A and B in order to simulate the number of cases when schools would have stayed open.

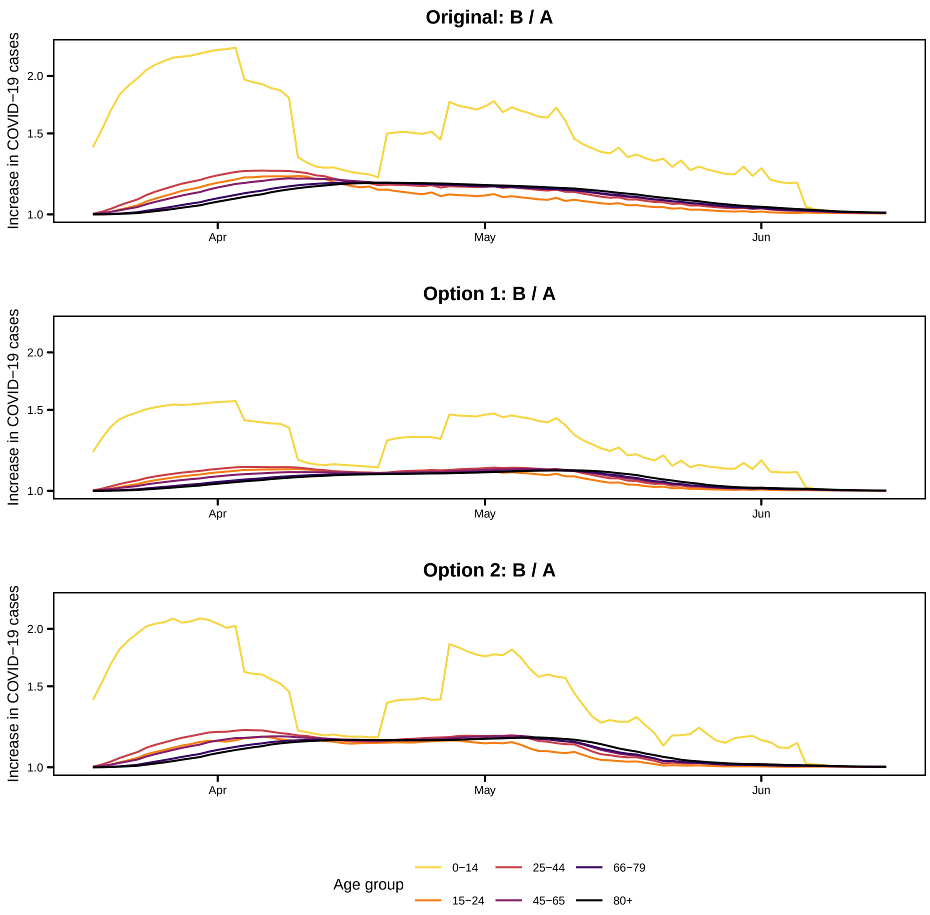


Figure S9: Ratio of the age group-specific path trajectories (B divided by A) throughout time

Both options dampen the number of cases as would be expected since the dominance of contacts in school settings (represented through $d_{s,t}$ is now lower. Option 1 has a lower number of expected cases than Option 2. Both have similar trickle effects to other age groups but the peak is lower for the school and working aged age groups (all ages up to 65) and later for the oldest age groups (ages 66 and above).

Table S5: Comparison between scenarios A (true measures) and B (schools open) for the original method

| Age | B - A | | | B / A | | |
|---|---|---|---|---|---|---|
| | $P_{10}$ | $P_{50}$ | $P_{90}$ | $P_{10}$ | $P_{50}$ | $P_{90}$ |
| 0-14 | 162.9 | 240.0 | 404 | 1.76 | 1.81 | 1.87 |
| 15-24 | 45.1 | 89.9 | 218 | 1.06 | 1.08 | 1.12 |
| 25-44 | 192.9 | 362.3 | 821 | 1.10 | 1.12 | 1.15 |
| 45-65 | 153.3 | 311.5 | 773 | 1.07 | 1.09 | 1.12 |
| 66-79 | 48.9 | 113.8 | 323 | 1.04 | 1.06 | 1.08 |
| 80+ | 37.3 | 90.5 | 271 | 1.04 | 1.05 | 1.07 |
| Total (summed) | 641.7 | 1,207.1 | 2,820 | 1.09 | 1.11 | 1.13 |

Table S6: Comparison between scenarios A (true measures) and B (schools open) for Option 1

| Age | B - A | | | B / A | | |
|---|---|---|---|---|---|---|
| | $P_{10}$ | $P_{50}$ | $P_{90}$ | $P_{10}$ | $P_{50}$ | $P_{90}$ |
| 0-14 | 114.3 | 229 | 701 | 1.31 | 1.36 | 1.40 |
| 15-24 | 45.4 | 137 | 605 | 1.04 | 1.06 | 1.07 |
| 25-44 | 171.0 | 477 | 2,014 | 1.06 | 1.07 | 1.08 |
| 45-65 | 155.6 | 497 | 2,334 | 1.04 | 1.05 | 1.06 |
| 66-79 | 64.5 | 248 | 1,301 | 1.03 | 1.04 | 1.05 |
| 80+ | 53.9 | 224 | 1,236 | 1.02 | 1.03 | 1.05 |
| Total (summed) | 604.9 | 1,815 | 8,188 | 1.05 | 1.06 | 1.07 |

Table S7: Comparison between scenarios A (true measures) and B (schools open) for Option 2

| Age | B - A | | | B / A | | |
|---|---|---|---|---|---|---|
| | $P_{10}$ | $P_{50}$ | $P_{90}$ | $P_{10}$ | $P_{50}$ | $P_{90}$ |
| 0-14 | 79.4 | 117.5 | 232 | 1.49 | 1.51 | 1.53 |
| 15-24 | 24.4 | 58.3 | 181 | 1.04 | 1.06 | 1.11 |
| 25-44 | 101.4 | 223.1 | 662 | 1.06 | 1.09 | 1.13 |
| 45-65 | 89.7 | 223.2 | 719 | 1.05 | 1.08 | 1.11 |
| 66-79 | 36.8 | 109.6 | 387 | 1.04 | 1.07 | 1.10 |
| 80+ | 29.0 | 90.6 | 328 | 1.04 | 1.07 | 1.10 |
| Total (summed) | 359.8 | 823.6 | 2,517 | 1.06 | 1.09 | 1.12 |

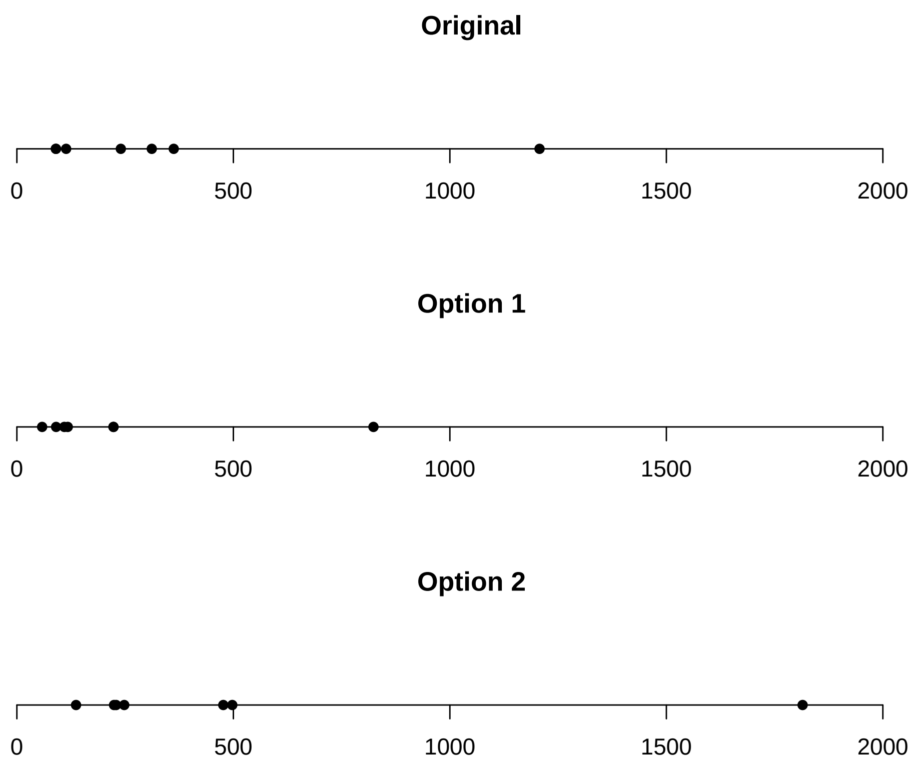


Figure S10: Visual comparison of the $P_{50}$ values for the three options

No difference is found in the ratios for scenario B so the relative measures are the same in this instance. The case counts are greater for Option 2 and lower for Option 1 (Figure S12).

##### 4.2 Scenario C

We also examine the predicted final of the epidemic when schools stay closed under the two alternative options for transmission weights. Now the difference in path trajectories is not as obvious, which means the differences seen for scenario B could also be an artefact of fewer cases early in the pandemic. The decrease in cases for June and July seems larger for the unadjusted weights which again is due to the dominance of contacts in school settings. Here the difference between Option 1 and Option 2 is still apparent with Option 1 having a smaller decrease and Option 2 showing a greater decrease.

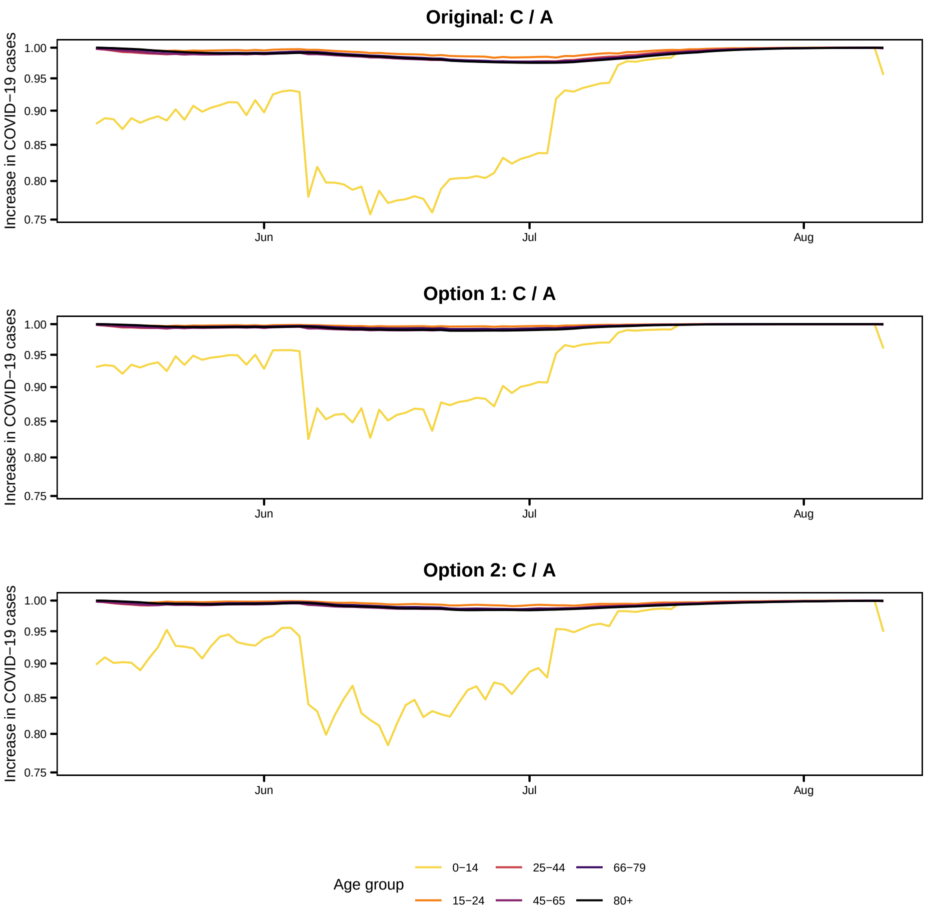


Figure S11: Ratio of the age group-specific trajectories (C divided by A) throughout time

#### 4.3 Discussion

The comparison of scenarios A and B as well as A and C for the various options showed a lot of differences for the adjusted versions against the original. The youngest age group remains the most visible in the path trajectories and the spread of

Table S8: Comparison between scenarios A (true measures) and C (schools closed) for the original method

| Age | C - A | | | C / A | | |
|---|---|---|---|---|---|---|
| | $P_{10}$ | $P_{50}$ | $P_{90}$ | $P_{10}$ | $P_{50}$ | $P_{90}$ |
| 0-14 | -27.5 | -20.85 | -16.98 | 0.915 | 0.922 | 0.928 |
| 15-24 | -16.6 | -9.38 | -5.98 | 0.990 | 0.994 | 0.995 |
| 25-44 | -64.4 | -36.03 | -23.15 | 0.985 | 0.989 | 0.991 |
| 45-65 | -55.2 | -30.08 | -18.96 | 0.983 | 0.988 | 0.990 |
| 66-79 | -13.5 | -6.92 | -4.05 | 0.984 | 0.988 | 0.991 |
| 80+ | -9.5 | -4.69 | -2.62 | 0.982 | 0.987 | 0.989 |
| Total (summed) | -186.0 | -107.83 | -71.69 | 0.983 | 0.987 | 0.990 |

Table S9: Comparison between scenarios A (true measures) and C (schools closed) for Option 1

| Age | C - A | | | C / A | | |
|---|---|---|---|---|---|---|
| | $P_{10}$ | $P_{50}$ | $P_{90}$ | $P_{10}$ | $P_{50}$ | $P_{90}$ |
| 0-14 | -14.29 | -11.81 | -10.19 | 0.957 | 0.959 | 0.961 |
| 15-24 | -8.81 | -5.16 | -3.48 | 0.995 | 0.996 | 0.997 |
| 25-44 | -35.16 | -20.76 | -14.12 | 0.992 | 0.994 | 0.995 |
| 45-65 | -31.28 | -17.76 | -11.73 | 0.991 | 0.993 | 0.994 |
| 66-79 | -9.24 | -4.83 | -2.93 | 0.991 | 0.994 | 0.995 |
| 80+ | -6.20 | -3.17 | -1.82 | 0.991 | 0.993 | 0.994 |
| Total (summed) | -104.77 | -63.34 | -44.47 | 0.991 | 0.993 | 0.994 |

Table S10: Comparison between scenarios A (true measures) and C (schools closed) for Option 2

| Age | C - A | | | C / A | | |
|---|---|---|---|---|---|---|
| | $P_{10}$ | $P_{50}$ | $P_{90}$ | $P_{10}$ | $P_{50}$ | $P_{90}$ |
| 0-14 | -8.889 | -8.124 | -7.522 | 0.963 | 0.965 | 0.966 |
| 15-24 | -1.589 | -1.244 | -1.009 | 0.999 | 0.999 | 0.999 |
| 25-44 | -6.902 | -5.590 | -4.700 | 0.997 | 0.998 | 0.998 |
| 45-65 | -5.499 | -4.409 | -3.678 | 0.997 | 0.997 | 0.997 |
| 66-79 | -1.036 | -0.758 | -0.580 | 0.998 | 0.998 | 0.998 |
| 80+ | -0.691 | -0.493 | -0.359 | 0.997 | 0.997 | 0.998 |
| Total (summed) | -24.524 | -20.604 | -17.903 | 0.996 | 0.997 | 0.997 |

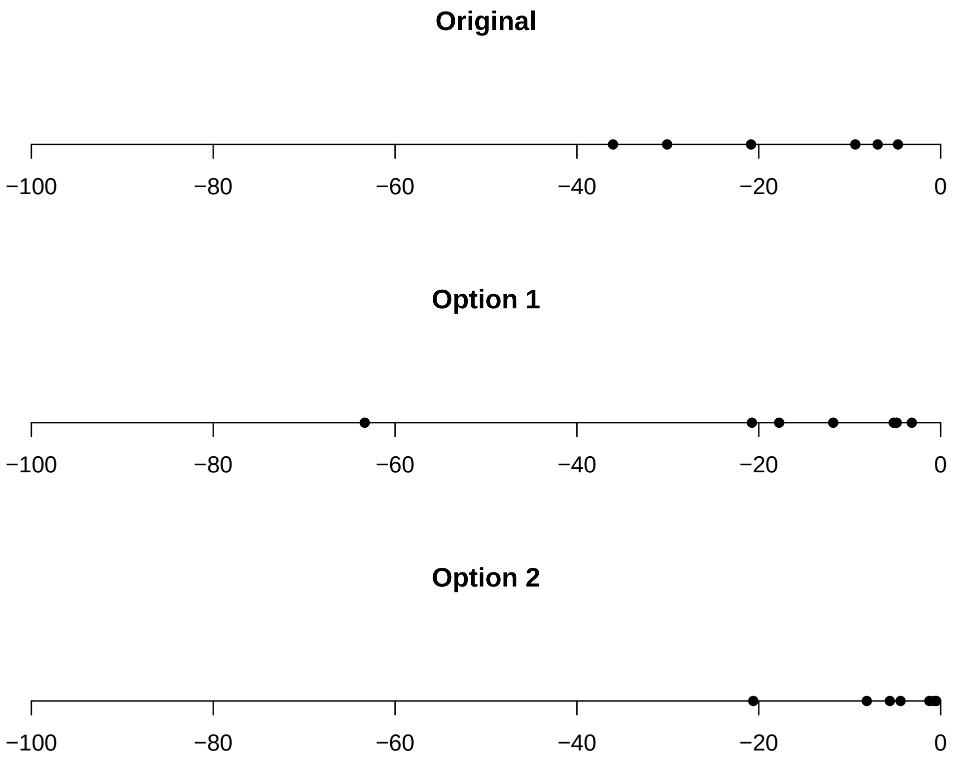


Figure S12: Visual comparison of the $P_{50}$ values for the three options

cases to other ages groups is more evident early on when schools are open (scenario B) than the reduction in cases later on when schools are closed (scenario C).

#### References
